# Effectiveness of COVID-19 vaccines against Omicron and Delta hospitalisation: test negative case-control study

**DOI:** 10.1101/2022.04.01.22273281

**Authors:** Julia Stowe, Nick Andrews, Freja Kirsebom, Mary Ramsay, Jamie Lopez Bernal

**Affiliations:** UK Health Security Agency, London, United Kingdom; NIHR Health Protection Research Unit in Vaccines and Immunisation, London School of Hygiene and Tropical Medicine, London, United Kingdom; NIHR Health Protection Research Unit in Respiratory Infections, Imperial College London, United Kingdom

**Keywords:** COVID-19, Omicron, Delta, Vaccine Effectiveness, Test Negative Case Control, Immunisation, Hospitalisation

## Abstract

**Background:** The omicron (B.1.1.529) variant has been associated with reduced vaccine effectiveness (VE) against infection and mild disease with rapid waning, even after a third dose, nevertheless omicron has also been associated with milder disease than previous variants. With previous variants protection against severe disease has been substantially higher than protection against infection.

**Methods:** We used a test-negative case–control design to estimate VE against hospitalisation with the omicron and delta variants using community and in hospital testing linked to hospital records. As a milder disease, there may be an increasing proportion of hospitalised individuals with Omicron as an incidental finding. We therefore investigated the impact of using more specific and more severe hospitalisation indicators on VE.

**Results:** Among 18-64 year olds using all Covid-19 cases admitted via emergency care VE after a booster peaked at 82.4% and dropped to 53.6% by 15+ weeks after the booster; using all admissions for >= 2 days stay with a respiratory code in the primary diagnostic field VE ranged from 90.9% down to 67.4%; further restricting to those on oxygen/ventilated/on intensive care VE ranged from 97.1% down to 75.9%. Among 65+ year olds the equivalent VE estimates were 92.4% down to 76.9%; 91.3% down to 85.3% and 95.8% down to 86.8%.

**Conclusions:** With generally milder disease seen with Omicron, in particular in younger adults, contamination of hospitalisations with incidental cases is likely to reduce VE estimates against hospitalisation. VE estimates improve and waning and waning is more limited when definitions of hospitalisation that are more specific to severe respiratory disease are used.

## Introduction

There has been a global increase in COVID-19 cases associated with the Omicron variant between November 2021 and March 2022.(1) Nevertheless surges in severe diseases, as indicated by hospitalisations, ICU admissions or deaths, have not matched those of previous waves of the pandemic.(2) A range of factors are likely to contribute to this divergence, including lower inherent severity of Omicron compared to previous variants, a greater proportion of the population with immunity from vaccination and/or prior infection, and sustained protection against severe disease.(3, 4)

Early data indicated a reduced neutralizing antibody response to the Omicron variant.(5-7) Real world studies have since found reduced effectiveness of COVID-19 vaccines against infection or mild disease with the Omicron variant.(8-10) Receipt of a booster dose improves protection, however, this appears to wane rapidly from the second month after vaccination.(8) Evidence on protection against severe disease is mixed with some studies suggesting substantially reduced effectiveness against hospitalisation compared to the Delta variant even with booster doses,(11, 12) whereas other studies suggest very high levels of effectiveness of over 90%.(9, 13, 14) There is currently limited data on the duration of protection against severe disease.

In this study, we assess the effectiveness of COVID-19 vaccines against hospitalisation in those testing positive by PCR for Omicron and Delta variants. In the past we have done this using symptomatic community tested cases subsequently hospitalised through emergency care for a non-accident reason within 2 weeks of their positive test with a test-negative case-control (TNCC) design.(15) This has yielded estimates of effectiveness of over 90% against Alpha and Delta variants. However, given that all individuals who are hospitalised for any reason in the UK are tested for COVID-19, and with the lower severity of Omicron and the high incidence, an increasing proportion of those hospitalised who also test positive may be hospitalised with COVID-19 as an incidental finding rather than hospitalised as a result of COVID-19. This would lead to underestimation of effectiveness against hospitalisation because the “with COVID-19” cases would be expected to have effectiveness similar to that seen against infection. To investigate this specificity of outcome issue we have obtained data on coded hospital discharges in those PCR tested including on primary diagnosis, length of stay, oxygen use, ventilation and admission to intensive care. Whilst these data are not as timely as using emergency care admission data, they allow identification of those more likely to admitted due to COVID-19.

## Methods

### Study Design

A test negative case control design was used to estimate vaccine effectiveness in those aged 18 years and over against hospitalisation following a PCR test for SARS-CoV-2. Cases were those testing positive and controls those testing negative by PCR. Effectiveness was assessed using a variety of hospitalisation end points designed to differentiate between hospitalisations likely to be because of COVID-19 and those that may be hospitalisation with COVID-19 but potentially due to another cause. Effectiveness against Omicron and Delta was assessed using periods in which these variants were circulating and using information on sequencing, genotyping and PCR s-gene target.

### Data Sources

#### COVID-19 Testing Data

PCR testing for SARS CoV-2 in England is undertaken by hospital and public health laboratories (Pillar 1), as well as by community testing (Pillar 2). Pillar 2 testing was available to anyone with symptoms consistent with COVID-19 (high temperature, new continuous cough, or loss or change in sense of smell or taste), anyone who was a contact of a confirmed case, care home staff and residents, and to those who tested positive using a lateral flow test (LFT). Pillar 1 testing is PCR testing in public health laboratories and NHS hospitals and was available for inpatients and others presenting to secondary care as well as health and care workers.

Data on all positive tests regardless of symptom status (PCR and LFT) and negative PCR tests from Pillar 2 from symptomatic individuals and all Pillar 1 tests with a sample date from 25 November 2020 to 10 March 2022 were identified. Where participants had a positive test within 14 days of another positive, the earliest PCR test was used, but sequencing, genotyping and S-gene target status from later tests was retained. Where tests were on the same day pillar 2 symptomatic tests were retained. Positive and negative tests within 90 days of a previous positive test and negative tests taken within 21 days after a positive test were excluded. Data were restricted to tests with a valid NHS number so linkage to the vaccination record could be carried out.

Classification of positive samples as Delta and Omicron variants was done using, in order of priority, whole genome sequencing, genotyping and S-gene target status. From sequencing Omicron is VOC-21NOV-01 or VUI-22JAN-01and Delta is VOC-21APR-02 or VUI-21OCT-01. S-gene target failure as well as genotyping and sequencing data could be used to identify Omicron from 29 November 2021 onward, however from 10 January 2022 Delta was very rare so all samples were assigned as Omicron if no sequencing or genotyping was done.(8) The Delta variant was classified using only sequencing, genotyping and S-gene target non-failure for the periods of 26 April 2021-23 May 2021 and 22 November 2021 to 3 January 2022, whilst for the period from 24 May to 21 November 2022, all samples were defined as Delta if no sequencing, genotyping or S-gene testing was done. Only those individuals where the variant was classified as Delta or Omicron were retained for analysis. The study period was therefore 26 April 2021 to 3 January 2022 for Delta and 22 November 2022 to2 February 2022 for Omicron.

#### Vaccination Data

These testing data were linked to the vaccination histories and demographic characteristics of the populations on 14 March 2022 using The National Immunisation Management System (NIMS) as previously described.(8, 16) Booster doses were identified as a third dose given at least 84 days after a second dose and administered after 13 September 2021. Individuals with vaccination histories outside the recommended schedules were excluded from the analysis (Figure 1).

**Figure 1:**
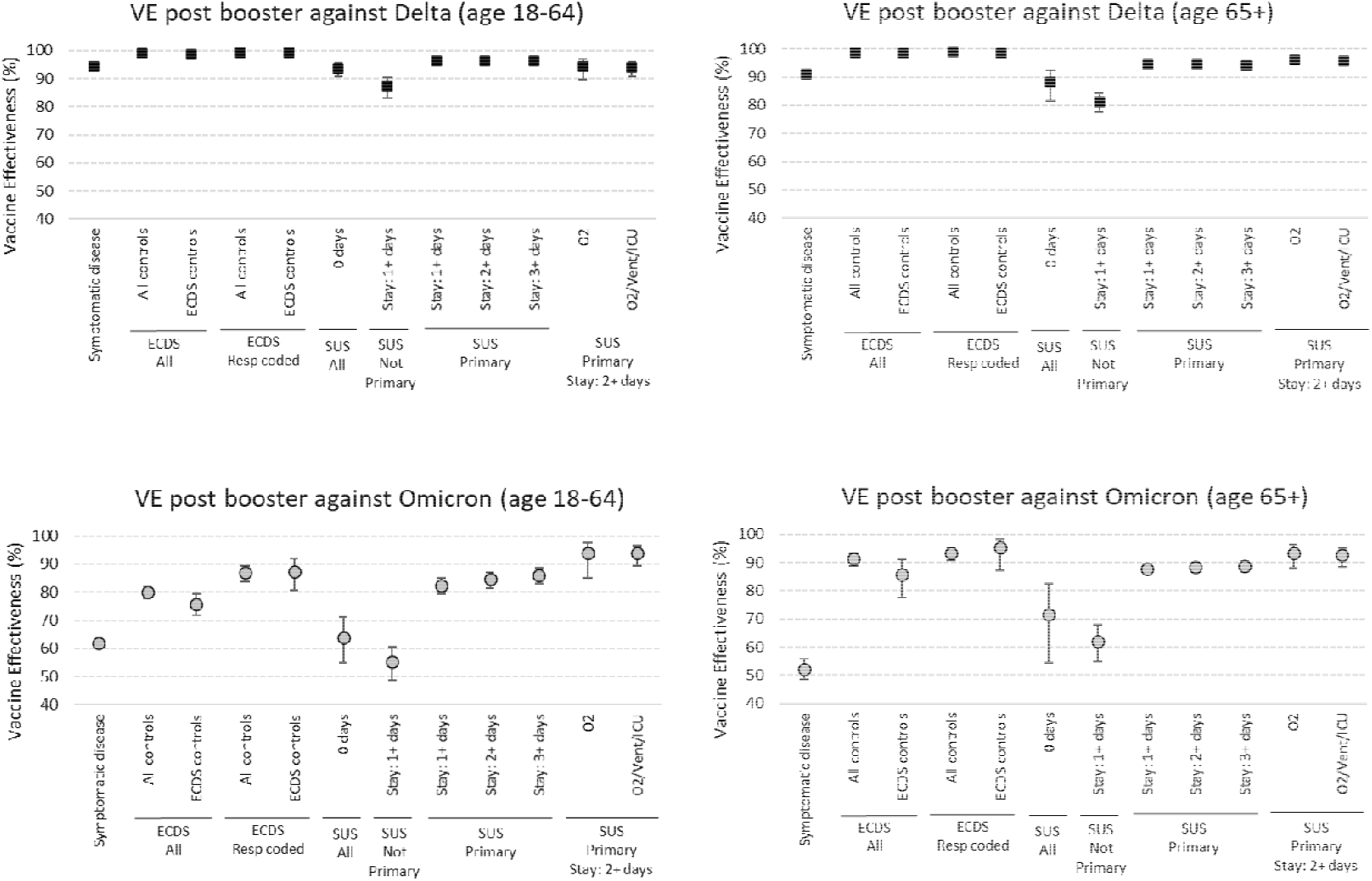
Vaccine effectiveness 7+ days after a booster dose against symptomatic disease and different hospitalisation outcomes by age group and variant

Testing data were linked to NIMS on 14 March 2022 using combinations of the unique individual National Health Service (NHS) number, date of birth, surname, first name, and postcode using deterministic linkage.

#### Emergency Care Hospital Admission Data

Emergency Care hospital admissions from the Emergency Care Dataset (ECDS), which includes hospital admissions through emergency departments but not elective admissions, were linked using NHS number and date of birth to the testing data on 15 March 2022 to identify admissions within 14 days of a community test. Admissions due to an injury were excluded. Admissions were identified where the Emergency Care Destination code was either discharge to a ward, intensive care unit, coronary care unit, high dependency unit or where there was a date on which the decision to admit the patient was made. Admissions with the reason for attending emergency care being a SNOMED CT (Systematized Nomenclature of Medicine–Clinical Terms) coded acute respiratory illness (ARI) were flagged.(17)

#### Secondary Care Hospital Admission Data

Hospital inpatient admissions for a range of acute respiratory illnesses (ARI) were identified from the Secondary Uses Service (SUS). SUS is the national electronic database of hospital admissions that provides timely updates of ICD-10 codes for completed hospital stays for all NHS hospitals in England. Up to 24 ICD-10 diagnoses fields can be completed in SUS for each admission with the first diagnosis field indicating the primary reason for admission. Oxygen us and ventilation support was ascertained using the Classification of Interventions and Procedures (OPCS-4) codes (table S15). Intensive Care Unit (ICU) admission status was ascertained by the Main Specialty of the ward being Critical Care Medicine or the Treatment Function being Intensive Care Medicine. Length of stay was calculated as date of discharge – date of admission.

For the Pillar 2 samples admissions with a ICD-10 acute respiratory illness (ARI) discharge diagnosis, in any diagnosis field, were identified where the sample was taken 14 days before and up to 2 days after the day of admission. For the Pillar 1 samples, admissions with an ICD-10 coded ARI discharge diagnosis in any diagnosis field, were identified where the sample was taken 1 days before and up to 2 days after the admission. The data was restricted to tests up to 23 February 2022 to account for delays in the SUS data recording. Linkage to the testing data was carried out on 15 March 2022 using NHS number and date of birth. Where multiple admissions linked to the same sample date the first admission after the sample date was retained and episode length calculated by summing the stay length for each admission.

#### Control selection

A maximum of one negative test per person within each of the following approximate 3 month periods was selected at random: 26 April to 1 August 2021, 2 August 2021 to 21 November 2021, 22 November 2021 to 23 February 2022. For analyses that involved hospitalised controls any negative tests that led to a hospitalisation within 21 days of a previous hospital negative test were excluded.

### Statistical Analysis

Analysis was by logistic regression with the PCR test result as the dependent variable where those testing positives were cases and those testing negative controls. Vaccination status was included as an independent variable and effectiveness defined as 1-odds of vaccination in cases/odds of vaccination in controls. Vaccination status was defined using date of onset, or, if missing or in Pillar 1 where this was not obtained, date of sample. Status was stratified by dose and interval post vaccination at 0-27 and 28+ days post first dose, 0-13,14-174 and 175+ days post second dose and 0-6, 7-13,14-34, 35-69, 70-104, 105+ post booster dose. The analysis was also stratified by manufacturer (ChAdOx-1 or BNT162b2 2 dose priming, and BNT162b2 or mRNA-1273 boosting) and by variant (Delta and Omicron). The analyses done to assess effectiveness according to specificity of the hospitalisation are given in Table 1. The first analysis replicates those previously done for symptomatic infection and the following analyses using different criteria to allow comparison of emergency care and SUS data sources and to assess within SUS how VE changes based on whether the respiratory code is in the primary diagnostic field, the length of stay and the presence of codes for further interventions (oxygen, ventilator, ICU admission).

**Table 1:** analyses to assess vaccine effectiveness against symptomatic disease and hospitalisation end points.

| Analysis | Analysis name | Pillar of testing | Cases (test positive) | Controls (test negative) |
| --- | --- | --- | --- | --- |
| 1 | Symptomatic disease | 2 | Symptomatic infection | Symptomatic infection (same as cases) |
| 2 | ECDS All - All controls | 2 | Symptomatic ECDS admitted | Symptomatic infection |
| 3 | ECDS All - ECDS controls | 2 | Symptomatic ECDS admitted | Hospitalised (same as cases) |
| 4 | ECDS ARI coded – All controls | 2 | Symptomatic ECDS admitted with a Respiratory code | Symptomatic infection |
| 5 | ECDS ARI coded – ECDS controls | 2 | Symptomatic ECDS admitted with a Respiratory code | Hospitalised (same as cases) |
| 6 | SUS, 0 days | 1 & 2 | SUS admitted with length of stay = 0 | Hospitalised (same as cases) |
| 7 | SUS, Not Primary, 1+ days | 1 & 2 | SUS admitted with length of stay $\geq 1$ day and ICD code not primary | Hospitalised (same as cases) |
| 8-10 | SUS, Primary, stay 1+, 2+, 3+ | 1 & 2 | SUS admitted, ICD code primary $\geq 1$, $\geq 2$, $\geq 3$ days stay | Hospitalised (same as cases) |
| 11 | SUS, Primary, stay 2+, O2 | 1 & 2 | SUS admitted, $\geq 2$ days stay, oxygen use | Hospitalised (same as cases) |
| 12 | SUS, Primary, stay 2+, O2/Vent/ICU | 1 & 2 | SUS admitted, $\geq 2$ days stay, oxygen use or ventilation of ICU admission. | Hospitalised (same as cases) |

Vaccine effectiveness was adjusted in logistic regression models for age (5 year bands), sex, index of multiple deprivation (quintile), ethnic group, care home residence status (for age 65+), geographic region (NHS region), period (calendar week of test), health and social care worker status (for age <65), clinical risk group status (for age<65), clinically extremely vulnerable, severely immunosuppressed, and previously testing positive. All analyses were stratified by age 18-64 and 65+. For the vaccine manufacturer stratification only end points 2-5, 9 and 12 were considered and only for Omicron. Numbers were too small in those primed with mRNA-1273 to assess this schedule.

## Results

### Descriptive Characteristics

After linkage of testing data to hospitalised cases in ECDS or a ARI coded SUS episode and to the NIMS vaccination database, and selection of the Delta and Omicron assigned cases and the controls, the total number of tests in the study period was 409,985 of which 115,720 were cases and 294,265 controls. A total of 51,115 (44.2%) of these cases and 34,556(11.7%) of these controls had a pillar 2 test as the earliest test and of these 38,150 cases and 31,552 controls were symptomatic and included in the ECDS analysis. For the ECDS analyses where all symptomatic controls were used irrespective of hospitalisation the total number of controls included was 6,759,286 whilst the analysis to assess symptomatic vaccine effectiveness using the pillar 2 data included 27,256 cases along with these controls.

The characteristics of hospitalised cases and controls for the Omicron and Delta period analyses are shown in Table S1 (age 18 to 64) and Table S2 (age 65 years and over). Note that some controls contribute to both the Omicron and Delta analyses. Pillar 2 symptomatic ECDS admissions in cases are much lower than SUS admissions for those aged over 65, even when restricting to those with a 2 day stay and primary diagnostic field coded. This difference is less for age 18 to 64 and for Omicron (18 to 64). Of the SUS admissions the proportion with a recorded intervention (oxygen/ventilation/ICU) is higher for Delta cases (age 18 to 64: 20.8% ; age 65+: 21.2%) than Omicron cases (age 18 to 64:2.5% ; age 65+: 6.6%) and higher for cases than controls except for Omicron cases (2.5%) compared to controls (4.4%) for age 18 to 64. This indicates not only severity differences by variant but also that severity differences differ by age with particularly low severity in age 18 to 64 Omicron cases.

### Post booster effectiveness by outcome

Figure 1 and Table S3 summarises vaccine effectiveness at least 7 days post booster by age, variant and outcome. For Delta in those ages 18 to 64 and 65+ VE against symptomatic infection was just over 90%. For all Delta ECDS analyses VE was very high at over 98% irrespective of controls used or respiratory coding or age. For the Delta SUS analysis it is clear that those with 0 length of stay or not with a respiratory code in the primary field show lower VE, even lower than for symptomatic infection. The Delta SUS analyses with at least 2 days stay and a primary coding all show VE of over 93%. For Omicron results are much more variable. As previously seen VE against symptomatic infection in much lower than for Delta with point estimates of 62% (age 18 to 64) and 52% (age 65+). For those age 65+ VE against hospitalisation using ECDS data is 86-91% improving to 93-95% if respiratory coding is used and with little variation according to which control group is used. VE in SUS in this age group is much lower, and more similar to symptomatic infection VE if using 0 days length of stay or a non-primary respiratory diagnosis. With the more specific and severe SUS end points VE increases to over 88-93%% and is similar to that seen for ECDS. The picture in those aged 18 to 64 is more complex with ECDS data giving VE of 75-80% with an increase to 87% if respiratory coded. Using SUS data with 0 days admission or a non-primary respiratory code gives VE similar to that for symptomatic infection, but there is a large increase in VE as length of stay increases (to 89% VE) and with use of Oxygen (to 93% VE). Only when oxygen use forms part of the definition is the VE in those aged 18 to 64 similar to that seen in age 65+.

### Effectiveness by vaccine manufacture, dose and interval

Analysis using all outcomes by dose intervals post vaccination are summarised in Table 2 (for Omicron) and Table S4 (for Delta) with full details in tables S5-S10. They show the same general patterns as seen when concentrating on post booster effectiveness. With Delta, with almost all of the outcomes, limited waning is seen, in particular among 18 to 64 year olds. With Omicron waning is seen with the less specific and less severe outcomes, though this is less obvious with the more specific and more severe outcomes. More waning is seen among 18 to 64 year olds with all outcomes for Omicron.

**Table 2:** vaccine effectiveness against different hospitalisation outcomes with Omicron by dose and interval (all vaccines combined)

|  |  | P2s ECDS<br>(ECDS<br>controls) | P2s ECDS<br>ARI coded<br>(ECDS ARI<br>controls) | P1/2 SUS<br>0 days | P1/2 SUS<br>1+ days<br>Not<br>Primary | P1/2 SUS<br>1+ days<br>Primary | P1/2 SUS 2+<br>days Primary | P1/2 SUS 3+<br>days Primary | P1/2 SUS 2+<br>days Primary<br>& oxygen | P1/2 SUS 2+<br>days Primary<br>& oxygen,<br>ventilation,<br>or ICU |
| --- | --- | --- | --- | --- | --- | --- | --- | --- | --- | --- |
| Age 18-64 |  |  |  |  |  |  |  |  |  |  |
|  | Interval | VE (95% CI) | VE (95% CI) | VE (95% CI) | VE (95% CI) | VE (95% CI) | VE (95% CI) | VE (95% CI) | VE (95% CI) | VE (95% CI) |
| Dose 1 | 0-27 | 48.5 (12.3 to 69.7) | 76.0 (-1.8 to 94.3) | 21.9 (-89.7 to 67.9) |  | 40.3 (-18 to 69.8) | 36.2 (-33.9 to 69.6) | 40 (-40.4 to 74.4) |  |  |
|  | 28+ | 48.7 (32.8 to 60.8) | 75.0 (50.3 to 87.4) | 25.0 (-9.1 to 48.5) | 16.2 (-3.7 to 32.3) | 42.8 (26.3 to 55.5) | 44.1 (25.6 to 58.0) | 51.5 (33.2 to 64.8) | 87.5 (55.6 to 96.5) | 75.0 (42.4 to 89.1) |
| Dose 2 | 0-13 | 39.6 (-31.5 to 72.2) | 50.0 (-187.1 to 91.3) | 63.6 (-0.4 to 86.8) | 8.3 (-152.9 to 66.7) | 87.5 (59.5 to 96.1) | 88.9 (58.4 to 97.0) | 87.6 (53.7 to 96.7) |  |  |
|  | 14-174 | 54.7 (45.3 to 62.4) | 73.7 (56.9 to 84.0) | 46.9 (30.5 to 59.5) | 29.5 (15.1 to 41.5) | 71.6 (63.4 to 77.9) | 69.0 (58.1 to 77.0) | 72.7 (61.4 to 80.7) | 79.1 (-36.9 to 96.8) | 86.7 (63.6 to 95.1) |
|  | 175+ | 34.6 (21.7 to 45.4) | 46.5 (14.2 to 66.7) | 41.7 (25.3 to 54.5) | 17.8 (4.4 to 29.3) | 52.5 (43.3 to 60.1) | 56.1 (46.4 to 64.0) | 60.6 (50.7 to 68.5) | 80.5 (48.7 to 92.6) | 82.3 (67.7 to 90.3) |
| Booster | 0-6 | 63.9 (52.2 to 72.8) | 76.3 (50.5 to 88.7) | 62.6 (40.1 to 76.7) | 19.8 (-25.2 to 48.6) | 70.1 (51.8 to 81.4) | 74.3 (55.9 to 85.0) | 77.2 (59.5 to 87.2) | 77.2 (-73.6 to 97.0) | 90.7 (56.0 to 98.1) |
|  | 7-13 | 80.1 (73.5 to 85.1) | 91.4 (82.7 to 95.7) | 75.3 (61.1 to 84.3) | 58.5 (39.3 to 71.6) | 87.7 (79.9 to 92.5) | 90.9 (83.2 to 95.1) | 95.0 (89.1 to 97.7) |  |  |
|  | 14-34 | 82.4 (78.6 to 85.6) | 91.4 (85.5 to 94.9) | 72.7 (63.9 to 79.3) | 56.2 (47.3 to 63.7) | 87.8 (84.3 to 90.5) | 88.6 (84.9 to 91.5) | 89.8 (85.9 to 92.6) | 94.2 (76.6 to 98.6) | 97.1 (92.2 to 98.9) |
|  | 35-69 | 72.7 (67.2 to 77.2) | 86.2 (77.8 to 91.5) | 62.6 (52 to 70.9) | 56.6 (49.6 to 62.7) | 83.4 (80.0 to 86.2) | 85.8 (82.4 to 88.5) | 87.8 (84.3 to 90.4) | 93.9 (81.6 to 97.9) | 94.3 (88.9 to 97.1) |
|  | 70-104 | 66.9 (59.1 to 73.3) | 79.5 (64.5 to 88.1) | 44.8 (26.1 to 58.8) | 50.2 (40 to 58.7) | 76.3 (70.8 to 80.7) | 80.2 (74.9 to 84.4) | 80.4 (74.5 to 85) | 94.0 (78.8 to 98.3) | 89.9 (78.3 to 95.3) |
|  | 105+ | 53.6 (36.9 to 65.9) | 60.7 (14.7 to 81.9) | 11.7 (-36.5 to 42.9) | 50.9 (34.3 to 63.3) | 66.3 (53.6 to 75.5) | 67.4 (53.1 to 77.4) | 68.6 (52.3 to 79.4) | 80.4 (-36.3 to 97.2) | 75.9 (15.8 to 93.1) |

|  |  | Age 65+ |  |  |  |  |  |  |  |  |
| --- | --- | --- | --- | --- | --- | --- | --- | --- | --- | --- |
|  | Interval | VE (95% CI) | VE (95% CI) | VE (95% CI) | VE (95% CI) | VE (95% CI) | VE (95% CI) | VE (95% CI) | VE (95% CI) | VE (95% CI) |
| Dose 1 | 0-27 |  |  |  | 11.7 (-118.6 to 64.3) | 57.4 (-0.7 to 82.0) | 43.9 (-41.0 to 77.7) | 35.1 (-80.6 to 76.7) |  |  |
|  | 28+ |  |  | 67.5 (6.7 to 88.6) | 33.2 (5.2 to 53.0) | 52.3 (35.8 to 64.5) | 53.4 (36.3 to 65.9) | 57.6 (41.3 to 69.3) | 48.3 (-169.1 to 90.0) | 78.3 (43.7 to 91.7) |
|  | 14-174 | 77.8 (45.0 to 91.0) | 90.6 (46.8 to 98.4) | 55.1 (-131.8 to 91.3) | 66.5 (47.1 to 78.7) | 80.5 (72.2 to 86.3) | 82.3 (74.3 to 87.8) | 82.2 (73.7 to 87.9) | 87.7 (50.9 to 96.9) | 90.9 (72.6 to 97.0) |
|  | 175+ | 66.7 (43.4 to 80.4) | 79.7 (34.4 to 93.7) | 39.2 (-6.7 to 65.4) | 23.5 (6.4 to 37.5) | 58.4 (51.0 to 64.7) | 57.7 (49.6 to 64.4) | 57.8 (49.4 to 64.9) | 74.0 (47.6 to 87.1) | 73.4 (55.1 to 84.3) |
| Booster | 0-6 | 85.8 (61.5 to 94.7) | 97.3 (79.2 to 99.7) | 85.6 (6.4 to 97.8) | 38.9 (-2.6 to 63.6) | 78.5 (66.8 to 86.1) | 77.9 (65.3 to 85.9) | 77.9 (64.7 to 86.2) | 70.0 (-33.9 to 93.3) | 89.2 (63.1 to 96.8) |
|  | 7-13 | 92.3 (76.3 to 97.5) | 94.8 (-7.5 to 99.8) | 69.2 (-18.7 to 92.0) | 62.5 (40.7 to 76.4) | 82.2 (73.0 to 88.3) | 84.7 (76.0 to 90.2) | 84.5 (75.5 to 90.2) | 86.7 (-13.9 to 98.5) | 94.7 (71.6 to 99.0) |
|  | 14-34 | 92.4 (86.0 to 95.8) | 97.9 (92.6 to 99.4) | 87.4 (72.5 to 94.2) | 69.3 (60.8 to 76.0) | 91.3 (89.1 to 93.0) | 91.3 (89.1 to 93.1) | 91.4 (89.0 to 93.2) | 95.9 (89.0 to 98.4) | 95.8 (91.3 to 97.9) |
|  | 35-69 | 87.0 (79.2 to 91.8) | 95.3 (87.3 to 98.3) | 79.3 (65.7 to 87.5) | 67.2 (60.5 to 72.8) | 88.9 (87.1 to 90.6) | 89.3 (87.3 to 90.9) | 89.5 (87.5 to 91.2) | 93.9 (88.4 to 96.8) | 92.8 (88.4 to 95.6) |
|  | 70-104 | 84.0 (74.6 to 89.9) | 94.2 (84.0 to 97.9) | 67.2 (46.6 to 79.8) | 59.4 (51.5 to 66.0) | 87.6 (85.6 to 89.3) | 88.1 (86.1 to 89.9) | 88.6 (86.5 to 90.3) | 93.2 (87.5 to 96.2) | 92.5 (88.1 to 95.2) |
|  | 105+ | 76.9 (60.6 to 86.4) | 90.3 (67.8 to 97.1) | 59.0 (30.5 to 75.8) | 56.3 (46.9 to 64.0) | 84.1 (81.2 to 86.5) | 85.3 (82.4 to 87.6) | 86.4 (83.6 to 88.7) | 90.1 (79.7 to 95.2) | 86.8 (77.1 to 92.3) |

To assess effectiveness by manufacturer only the ECDS (all controls), ECDS respiratory coded (all controls), SUS primary code >=2 days stay and SUS, primary code >=2 days stay and Oxygen/ventilation/ICU end points were considered, and only for Omicron since Delta VE varies less by end point and results have been previously published.(18) These end points were chosen to be the same ECDS end point used in past analyses and to use the more specific SUS end points. Figure 2 shows the ECDS analysis with lower VE in those aged 18-64 and waning post booster, more so for the BNT162b2 booster where there is a longer follow-up where VE declines to 38% for those primed with ChAdOx-1. For those aged 65+ ECDS VE is higher at over 90% up to 14 weeks post booster irrespective or priming vaccine or the booster received and remaining over 80% from 15+ weeks after the booster. In this age the waned 2 dose VE is higher for BNT162b2 (76%) than ChAdOx-1 (56%). ECDS results with respiratory coding show generally higher VE with similar patterns (Tables S5-S8). Figure 3 shows the SUS results and shows VE above 80% in almost all vaccination combinations and post booster periods. For those aged over 65+ the SUS data suggest little evidence of waning, whilst in those aged 18-64 VE declines to around 66-69% 15+ weeks after a BNT162b2 boost. Within each interval, VE is similar for both ChAdOx-1 and BNT162b2 primed individuals and also for BNT162b2 and mRNA-1273 boosted individuals.

**Figure 2:**
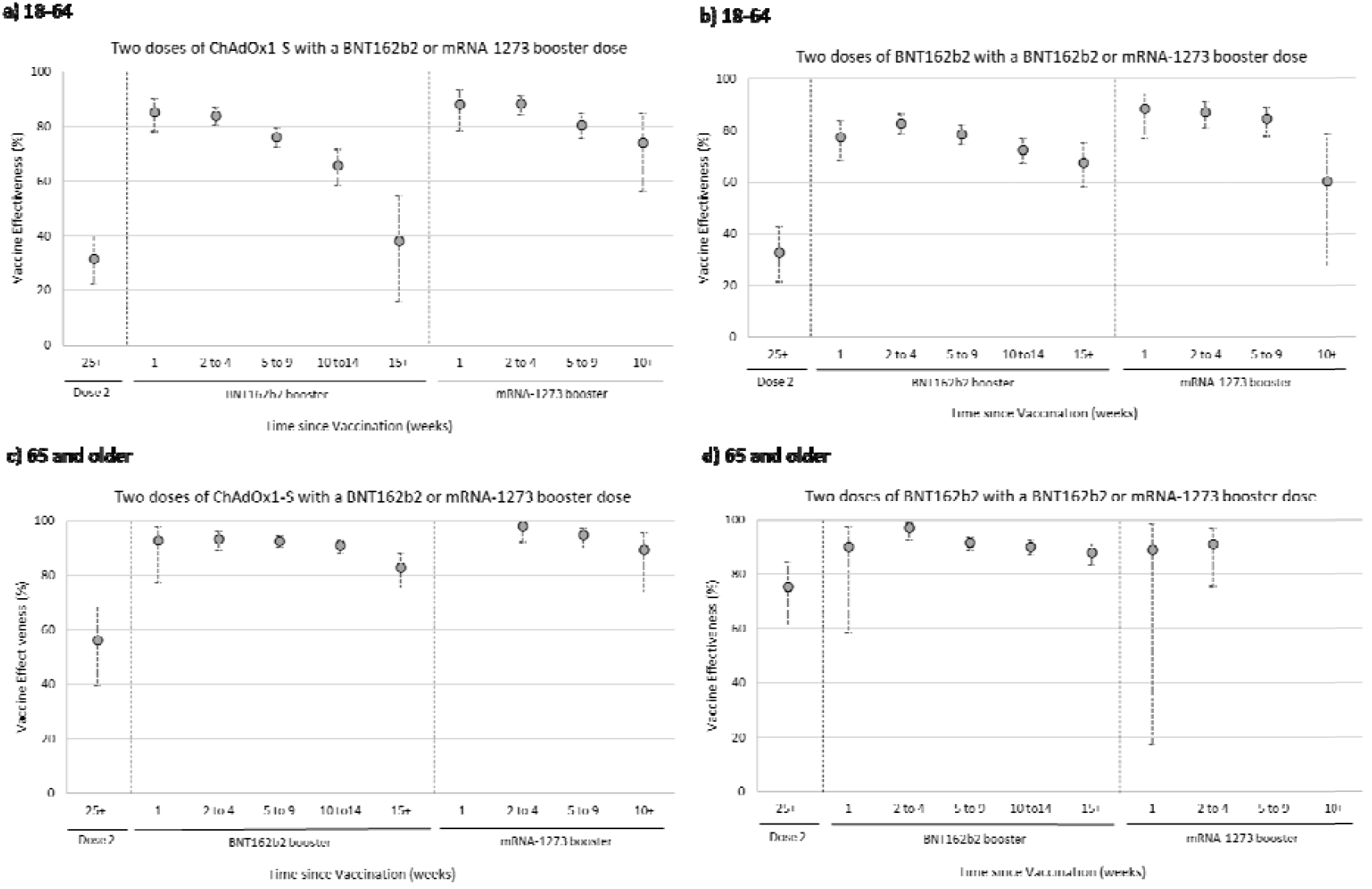
Vaccine effectiveness against hospitalisations using ECDS by age group and manufacturer (all symptomatic controls, Omicron only)

**Figure 3:**
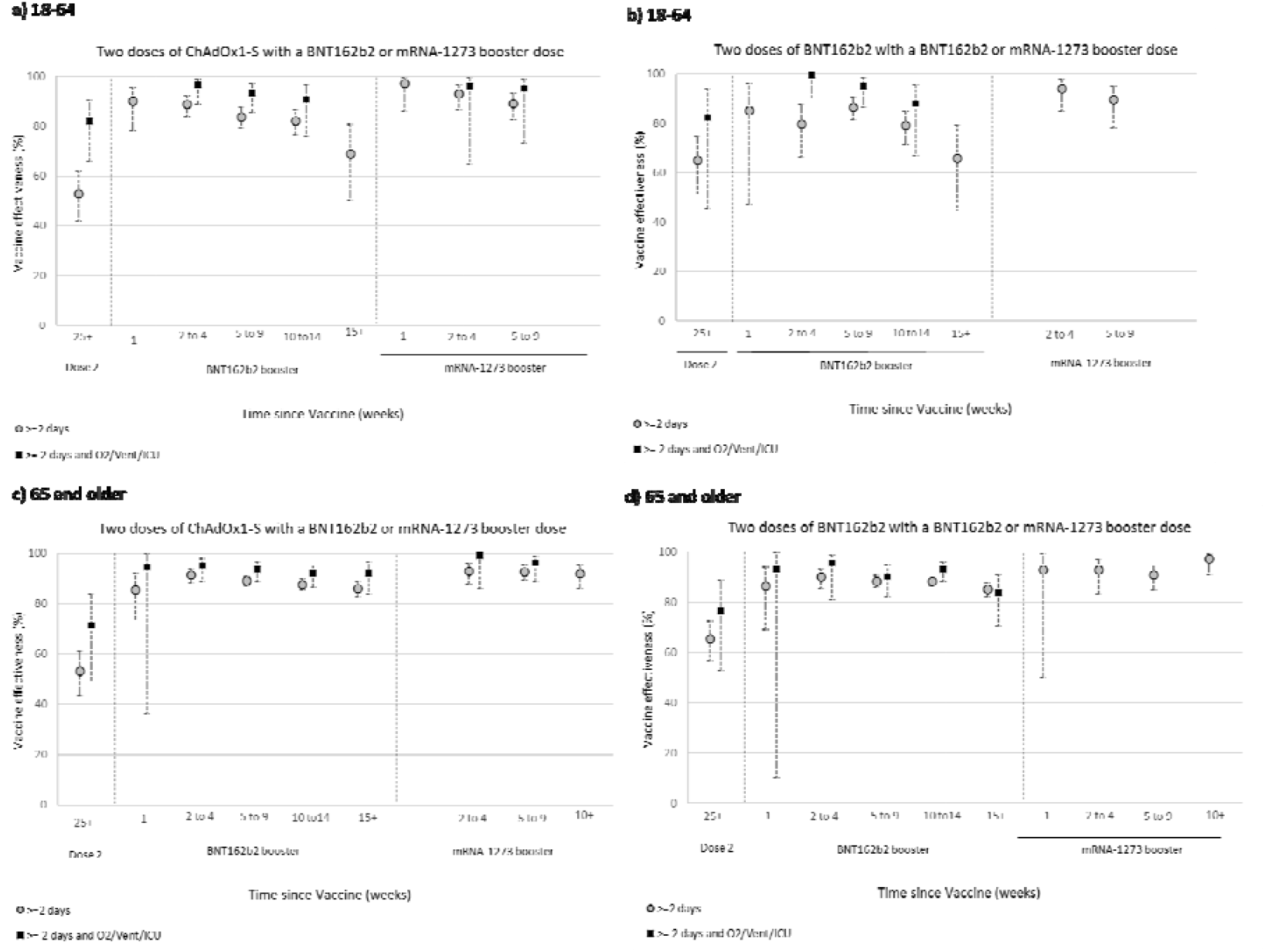
Vaccine effectiveness against hospitalisations >=2 days and >=2 days and on oxygen/ventilated/on ICU using SUS by age group and manufacturer (all symptomatic controls, Omicron only)

## Discussion

The results of this study demonstrate that assessment and interpretation of COVID-19 vaccine effectiveness against hospitalisation has become more complicated since the milder Omicron variant has become dominant. When the disease is milder a higher proportion of hospitalisations are likely to have COVID as an incidental finding rather than the cause of hospitalisation. This is the case for Omicron compared to Delta and for younger adults compared to older adults. Contamination of hospitalisations with these ‘incidental’ cases appears to result in lower vaccine effectiveness estimates against hospitalisation that are likely more reflective of vaccine effectiveness against infection. Vaccine effectiveness estimates improve and waning is more limited when definitions of hospitalisation that are more specific to severe respiratory disease are used.

For the Delta variant we found that VE was fairly robust using the emergency care admissions or SUS coded hospital discharges as long as the SUS discharge code was in the primary field and the admission length at least one overnight stay. The results suggest that for Delta a high proportion of these admissions are likely to be truly related to COVID-19 so that the VE measure is truly against a more severe disease. Furthermore, for Delta, contamination of the hospitalised cases with cases not hospitalised due to COVID will cause less bias for VE because symptomatic VE post booster is high. For the Omicron variant VE was also high and fairly robust to the case definition in those aged 65 and over, although it did increase when using respiratory coded ECDS admissions or when restricting to SUS cases with oxygen/ventilation or ICU. This age group also had the longest available follow-up post booster and largest numbers to look at VE by specific vaccine with these results showing similar VE by schedule post booster and with VE remaining high to 15+ weeks after the booster. In those aged 18-64 VE was lower at below 90% unless additional interventions (oxygen, ventilation or ICU) were included. VE against Omicron was particularly low and similar to symptomatic disease VE if using those without a primary respiratory code or admitted and discharged on the same day. This suggests these cases may be asymptomatically identified cases from screening of all hospitalised patients. When assessing VE against Omicron it is therefore not sufficient to just identify hospitalisation through routine hospital datasets without using more detailed data on diagnostic codes, length of stay and interventions. In previous reports we have given VE against hospitalisation through ECDS (all ages) and this has suggested declines by time since booster,(19) but this current analysis indicates how this is likely to be due, at least in part, to many of these hospitalisations not being due to COVID-19 leading to the estimates mirroring the declines seen against symptomatic infection.(8) Using admissions of at least 2 days with a respiratory code in the primary diagnostic field VE in both age groups started at around 91% soon after the booster, dropping to around 67% by 15+ weeks in 18 to 64 year olds and 85% in 65+ year olds. Among those on oxygen VE went from around 94% down to 80% in 18 to 64 year olds and 96% down to 90% in 65+ year olds. The lower VE and more notable waning among 18 to 64 year olds suggests that even with these more specific and more severe endpoints, there are likely to be a significant number of admissions where COVID is not the primary cause of their hospitalisation. Furthermore, among the 18 to 64 year olds, those who first became eligible for vaccination, and thus have the longest follow-up, are those in clinical risk groups, including immunosuppressed individuals – this is likely to contribute to the greater apparent waning in the last follow-up period.

Our findings may go some way towards explaining the differing findings among existing studies of vaccine effectiveness against severe disease with the Omicron variant. For example, Abu-Raddad et al found dose 3 vaccine effectiveness of 76.5% (95%CI, 55.9% to 87.5%) against Covid-19 related hospitalisation or death, which is lower than many other estimates.(11) This may be related to the fact that the study was dominated by under 60 year olds, who in general, are likely to have milder disease. Other studies where VE estimates after 3 doses were over 90% have included older cohorts or have used physician manual review of medical notes to confirm the presence of severe COVID-19 symptoms.(13, 14) We only identified one study that had stratified by period after a booster dose – Thompson et al found VE of 91% in the first 2 months following a third dose and 78%>= 4 months after the third dose – this is similar to our findings in 18 to 64 year olds with some of the outcomes, though generally more waning than that we observed in 65+ year olds.(14)

Limitations and advantages of the test negative case control design have been previously described.(8, 18, 20) One of the biggest limitations of this specific study is that in relies on hospital coded data which may have coding errors or not have interventions coded when they were used (e.g. oxygen use). A study where data are collected prospectively on cases using reporting forms or detailed case note review could avoid this misclassification bias, but is much more challenging to do with sufficiently large numbers.(21) One potential limitation for the TNCC design when looking at severe disease in controls is test sensitivity when a large proportion of those tested are truly positive. This, however, is more likely to affect Delta than Omicron analyses (as Delta is more severe) and is one of the reasons, along with study power, that in past analyses we have chosen to use all symptomatic pillar 2 controls for hospitalised COVID-19 VE. The analyses in this study do show slightly higher VE when using hospitalised pillar 2 controls which may be due to this bias, but which may also be due to residual confounding from using all controls because it is necessary to adjust for factors related to risk of hospitalisation. Another limitation is that we have not done a formal validation on cases using more detailed case note review to show that those with short stays and coding not in primary fields are less likely to be admitted due to COVID-19. Examining differences in VE by vaccine is particularly challenging given differences in the populations that have received either vaccine. For example, those that received ChAdOx1-S as the primary course are more likely to be in clinical risk group, particularly among younger age groups. Similarly, those in the youngest age groups that were vaccinated earliest are likely to be in clinical risk groups. While adjustments are made for age and clinical risk group, there is likely to be residual confounding.

This study has the advantage of having reasonably precise VE estimates due to the extensive testing that has been ongoing in the UK in both the community and hospitals. From the end of March 2022, however, community testing will be restricted to those in certain clinical risk groups. Although this will impact on assessment of VE against symptomatic infection, continuation of hospital testing will still allow assessment of VE against hospitalised COVID-19. This will be important to identify waning of the booster doses and the effectiveness of second spring boosters in those aged 75 and over.(22) It will also be important when new variants emerge to identify immune escape and its impact on more severe disease.

In conclusion, we found high levels of booster VE against hospitalisation with the Omicron variant, in particular among older adults who are at greatest risk, and against more severe end points. Nevertheless, there is evidence of limited waning from 3-4 months after a booster dose. Care should be taken in comparison of VE against hospitalisation across different studies due to the impact of using different outcome definitions.

## Data Availability

All data produced in the present work are contained in the manuscript. Data cannot be made publicly available for ethical and legal reasons, i.e. public availability would compromise patient
confidentiality as data tables list single counts of individuals rather than aggregated
data

## Acknowledgements

We thank the UK Health Security Agency (UKHSA) Covid-19 Data Science Team, UKHSA Outbreak Surveillance Team, NHS England, NHS Digital, and NHS Test and Trace for their roles in developing and managing the COVID-19 testing, variant identification and vaccination systems and datasets as well as reporting NHS vaccinators, NHS laboratories, UKHSA laboratories, and lighthouse laboratories; and we thank the Wellcome Sanger Institute and other laboratories involved in whole genome sequencing of COVID-19 samples; and we thank the Joint Committee on Vaccination and Immunisation and the UK Variant Technical Group for advice and feedback in developing this study.

## Ethics

Surveillance of coronavirus disease 2019 (Covid-19) testing and vaccination is undertaken under Regulation 3 of the Health Service (Control of Patient Information) Regulations 2002 to collect confidential patient information (www.legislation.gov.uk/uksi/2002/1438/regulation/3/made. opens in new tab) under Sections 3(i) (a) to (c), 3(i)(d) (i) and (ii), and 3. The study protocol was subject to an internal review by the Public Health England Research Ethics and Governance Group and was found to be fully compliant with all regulatory requirements. Given that no regulatory issues were identified and that ethics review is not a requirement for this type of work, it was decided that a full ethics review would not be necessary.

## References

1. World Health Organization. COVID-19 Weekly Epidemiological Update (Edition 82, published 8 March 2022): World Health Organization; 2022 [updated 08/03/2022. Available from: https://www.who.int/publications/m/item/weekly-epidemiological-update-on-covid-198-march-2022.

2. UK Health Security Agency. National flu and COVID-19 surveillance graphs: 10 March 2022 (week 10) 2020 [Available from: https://www.gov.uk/government/statistics/national-flu-and-covid-19-surveillance-reports-2021-to-2022-season.

3. Nyberg T, Ferguson NM, Nash SG, Webster HH, Flaxman S, Andrews N, et al. Comparative Analysis of the Risks of Hospitalisation and Death Associated with SARS-CoV-2 Omicron (B. 1.1. 529) and Delta (B. 1.617. 2) Variants in England. 2022.

4. Madhi SA, Kwatra G, Myers JE, Jassat W, Dhar N, Mukendi CK, et al. Population Immunity and Covid-19 Severity with Omicron Variant in South Africa. New England Journal of Medicine 2022.

5. Cele S, Jackson L, Khan K, Khoury D, Moyo-Gwete T, Tegally H, et al. SARS-CoV-2 Omicron has extensive but incomplete escape of Pfizer BNT162b2 elicited neutralization and requires ACE2 for infection. medRxiv. 2021:2021.12.08.21267417.

6. Wilhelm A, Widera M, Grikscheit K, Toptan T, Schenk B, Pallas C, et al. Reduced Neutralization of SARS-CoV-2 Omicron Variant by Vaccine Sera and monoclonal antibodies. medRxiv. 2021:2021.12.07.21267432.

7. Dejnirattisai W, Huo J, Zhou D, Zahradník J, Supasa P, Liu C, et al. Omicron-B.1.1.529 leads to widespread escape from neutralizing antibody responses. bioRxiv. 2021:2021.12.03.471045.

8. Andrews N, Stowe J, Kirsebom F, Toffa S, Rickeard T, Gallagher E, et al. Covid-19 Vaccine Effectiveness against the Omicron (B.1.1.529) Variant. New England Journal of Medicine. 2022.

9. Buchan SA, Chung H, Brown KA, Austin PC, Fell DB, Gubbay JB, et al. Effectiveness of COVID-19 vaccines against Omicron or Delta symptomatic infection and severe outcomes. medRxiv. 2022:2021.12.30.21268565.

10. Hansen CH, Schelde AB, Moustsen-Helm IR, Emborg H-D, Krause TG, Mølbak K, et al. Vaccine effectiveness against SARS-CoV-2 infection with the Omicron or Delta variants following a two-dose or booster BNT162b2 or mRNA-1273 vaccination series: A Danish cohort study. medRxiv. 2021:2021.12.20.21267966.

11. Abu-Raddad LJ, Chemaitelly H, Ayoub HH, AlMukdad S, Yassine HM, Al-Khatib HA, et al. Effect of mRNA Vaccine Boosters against SARS-CoV-2 Omicron Infection in Qatar. New England Journal of Medicine 2022.

12. Collie S, Champion J, Moultrie H, Bekker L-G, Gray G. Effectiveness of BNT162b2 Vaccine against Omicron Variant in South Africa. New England Journal of Medicine. 2021;386(5):494–6.

13. Tseng HF, Ackerson BK, Luo Y, Sy LS, Talarico CA, Tian Y, et al. Effectiveness of mRNA-1273 against SARS-CoV-2 omicron and delta variants. medRxiv. 2022:2022.01.07.22268919.

14. Thompson MG, Natarajan K, Irving SA, Rowley EA, Griggs EP, Gaglani M, et al. Effectiveness of a third dose of mRNA vaccines against COVID-19–associated emergency department and urgent care encounters and hospitalizations among adults during periods of Delta and Omicron variant predominance—VISION Network, 10 States, August 2021–January 2022 2022.

15. Andrews N, Tessier E, Stowe J, Gower C, Kirsebom F, Simmons R, et al. Duration of Protection against Mild and Severe Disease by Covid-19 Vaccines. New England Journal of Medicine. 2022.

16. NHS England. National COVID-19 and Flu Vaccination Programmes [Available from: https://www.england.nhs.uk/contact-us/privacy-notice/national-flu-vaccination-programme/.

17. UK Health Security Agency. Emergency department: weekly bulletins for 2021 2021 [Available from: https://www.gov.uk/government/publications/emergency-department-weekly-bulletins-for-2021.

18. Lopez Bernal J, Andrews N, Gower C, Gallagher E, Simmons R, Thelwall S, et al. Effectiveness of Covid-19 Vaccines against the B.1.617.2 (Delta) Variant. New England Journal of Medicine. 2021;385(7):585–94.

19. UK Health Security Agency. COVID-19 vaccine surveillance report Week 8 - 24 February 2022 2022 [Available from: https://assets.publishing.service.gov.uk/government/uploads/system/uploads/attachment_data/file/1057599/Vaccine_surveillance_report_-_week-8.pdf.

20. Lopez Bernal J, Andrews N, Gower C, Robertson C, Stowe J, Tessier E, et al. Effectiveness of the Pfizer-BioNTech and Oxford-AstraZeneca vaccines on covid-19 related symptoms, hospital admissions, and mortality in older adults in England: test negative case-control study. BMJ. 2021;373:n1088.

21. Hyams C, Marlow R, Maseko Z, King J, Ward L, Fox K, et al. Effectiveness of BNT162b2 and ChAdOx1 nCoV-19 COVID-19 vaccination at preventing hospitalisations in people aged at least 80 years: a test-negative, case-control study. The Lancet Infectious diseases. 2021.

22. Joint Committee on Vaccination and Immunisation. Joint Committee on Vaccination and Immunisation (JCVI) statement on COVID-19 vaccinations in 2022: 21 February 2022 2022 [Available from: https://www.gov.uk/government/publications/joint-committee-on-vaccination-and-immunisation-statement-on-covid-19-vaccinations-in-2022/joint-committee-on-vaccination-and-immunisation-jcvi-statement-on-covid-19-vaccinations-in-2022-21-february-2022.

