## Supplementary Tables for "Effectiveness of COVID-19 vaccines against Omicron and Delta hospitalisation: test negative case-control study"

[Table S5: Vaccine effectiveness against hospital admissions from emergency care (ECDS) within 14 days of the test date by the Delta and Omicron variants in symptomatic individuals 18 to 64 years of age](#_Toc98883738)…..11

[Table S6: Vaccine effectiveness against hospital admissions from emergency care (ECDS) within 14 days of the test date by the Delta and Omicron variants in symptomatic individuals 18 to 64 years of age using all controls	1](#_Toc98883739)2

[Table S7: Vaccine effectiveness against hospital admissions from emergency care (ECDS) within 14 days of the test date by the Delta and Omicron variants in symptomatic individuals 65 years of age and older….1](#_Toc98883740)3

[Table S8: Vaccine effectiveness against hospital admissions from emergency care (ECDS) within 14 days of the test date by the Delta and Omicron variants in symptomatic individuals 18 to 64 years of age using all controls	1](#_Toc98883741)4

[Table S9: Vaccine effectiveness using secondary care hospital admission data (SUS) by the Delta and Omicron variants in individuals 18-64 years of age	1](#_Toc98883742)5

[Table S10: Vaccine effectiveness using secondary care hospital admission data (SUS) by the Delta and Omicron variants in individuals 65 years of age and older	1](#_Toc98883743)8

[Table S11: Vaccine effectiveness by manufacturer against hospital admissions from emergency care (ECDS) within 14 days of the test date by the Omicron variant in symptomatic individuals 18 to 64 years of age	2](#_Toc98883744)1

[Table S12: Vaccine effectiveness by manufacturer against hospital admissions from emergency care (ECDS) within 14 days of the test date by the Omicron variant in symptomatic individuals 65 years of age and older	2](#_Toc98883745)2

[Table S13: Vaccine effectiveness using secondary care hospital admission data (SUS) by manufacturer for the Omicron variant in individuals 18-64 years of age	2](#_Toc98883746)4

[Table S14: Vaccine effectiveness using secondary care hospital admission data (SUS) by manufacturer for the Omicron variant in individuals 65 years of age and older	…………2](#_Toc98883747)5

Table S15: SUS ICD10 and OPCS code lists …………………….……………………………………………….………………………26

Table S16: Flow chart demonstrating the selection of the data in the study…………………………….…………….27

### Table S1. Descriptive characteristics of positive and negative test results in individuals 18-64 years of age tested for SARS-CoV-2 in England for the study population

|  |  |  | **Delta** | | | | **Omicron** | | | |
| --- | --- | --- | --- | --- | --- | --- | --- | --- | --- | --- |
|  |  |  | **Positive** | | **Negative** | | **Positive** | | **Negative** | |
|  |  |  | **n** | **%** | **n** | % | **n** | **%** | **n** | % |
| **Category** |  |  | **52,155** |  | **81,821** |  | **19,790** |  | **22,548** |  |
| **Vaccination Status after vaccine** | **Manufacturer (primary course)** | **Dose** |  |  |  |  |  |  |  |  |
|  | Unvaccinated | | 29,877 | 57.3% | 13,942 | 17.0% | 5,136 | 26.0% | 2,822 | 12.5% |
|  | ChAdOx1-S | Dose 1 | 1,825 | 3.5% | 6,180 | 7.6% | 409 | 2.1% | 447 | 2.0% |
|  |  | Dose 2 | 13,385 | 25.7% | 31027 | 37.9% | 2401 | 12.1% | 3930 | 17.4% |
|  |  | Booster (any) | 368 | 0.7% | 5210 | 6.4% | 4578 | 23.1% | 7310 | 32.4% |
|  | BNT162b2 | Dose 1 | 2672 | 5.1% | 4014 | 4.9% | 761 | 3.8% | 581 | 2.6% |
|  |  | Dose 2 | 3548 | 6.8% | 16820 | 20.6% | 2473 | 12.5% | 2807 | 12.4% |
|  |  | Booster (any) | 235 | 0.5% | 3637 | 4.4% | 3708 | 18.7% | 4280 | 19.0% |
|  | mRNA-1273 | Dose 1 | 187 | 0.4% | 387 | 0.5% | 66 | 0.3% | 73 | 0.3% |
|  |  | Dose 2 | 58 | 0.1% | 572 | 0.7% | 154 | 0.8% | 217 | 1.0% |
|  |  | Booster (any) | 0 | 0.0% | 32 | 0.0% | 104 | 0.5% | 81 | 0.4% |
| **Outcomes** | ECDS admissions -pillar 2 symptomatic | | 17,232 | 100.0% | 19,461 | 100.0% | 3,519 | 100.0% | 6,432 | 100.0% |
|  | ECDS admissions -pillar 2 symptomatic with respiratory code | | 9,618 | 55.8% | 3,070 | 15.8% | 782 | 22.2% | 973 | 15.1% |
|  | Secondary Uses Service Respiratory coded admissions (SUS) | | 47,550 | 100.0% | 63,781 | 100.0% | 17,075 | 100.0% | 16,527 | 100.0% |
|  | Secondary Uses Service Respiratory coded admissions (SUS) with 0 days length of stay | | 5,766 | 12.1% | 7,808 | 12.2% | 5,626 | 32.9% | 2,333 | 14.1% |
|  | SUS with respiratory code but NOT in primary diagnosis field with at least 1 day stay | | 9,395 | 19.8% | 29,601 | 46.4% | 7,469 | 43.7% | 7,308 | 44.2% |
|  | SUS with respiratory code as the primary diagnosis and length of stay >=1 day | | 32,389 | 68.1% | 26,372 | 41.3% | 3,980 | 23.3% | 6,886 | 41.7% |
|  | SUS with respiratory code as the primary diagnosis and length of stay >=2 days | | 28,537 | 60.0% | 21,834 | 34.2% | 2,827 | 16.6% | 5,568 | 33.7% |
|  | SUS with respiratory code as the primary diagnosis and length of stay >=3 days | | 25,163 | 52.9% | 18,193 | 28.5% | 2,110 | 12.4% | 4,606 | 27.9% |
|  | SUS >=2 days stay and primary diagnosis with oxygen use | | 5,866 | 12.3% | 1,385 | 2.2% | 222 | 1.3% | 336 | 2.0% |
|  | SUS >=2 days stay and primary diagnosis with oxygen use or ventilation used or in ICU | | 9,898 | 20.8% | 2,980 | 4.7% | 424 | 2.5% | 729 | 4.4% |
| **Testing route** | Pillar 1 |  | 24,007 | 46.0% | 58,249 | 71.2% | 10,457 | 52.8% | 15,055 | 66.8% |
|  | Pillar 2 |  | 28,148 | 54.0% | 23,572 | 28.8% | 9,333 | 47.2% | 7,493 | 33.2% |
| **Gender** | Female |  | 27,731 | 53.2% | 43,406 | 53.0% | 13,042 | 65.9% | 12,064 | 53.5% |
|  | Male |  | 24,338 | 46.7% | 37,772 | 46.2% | 6,717 | 33.9% | 10,317 | 45.8% |
|  | Missing |  | 86 | 0.2% | 643 | 0.8% | 31 | 0.2% | 167 | 0.7% |
| **Age** | 18-19 |  | 1,024 | 2.0% | 1,893 | 2.3% | 545 | 2.8% | 509 | 2.3% |
|  | 20-24 |  | 3,507 | 6.7% | 4,690 | 5.7% | 1838 | 9.3% | 1219 | 5.4% |
|  | 25-29 |  | 5,288 | 10.1% | 5,515 | 6.7% | 2586 | 13.1% | 1,588 | 7.0% |
|  | 30-34 |  | 6,479 | 12.4% | 6,607 | 8.1% | 2872 | 14.5% | 1,908 | 8.5% |
|  | 35-39 |  | 6,276 | 12.0% | 6,683 | 8.2% | 2263 | 11.4% | 1,960 | 8.7% |
|  | 40-44 |  | 5,797 | 11.1% | 6,789 | 8.3% | 2000 | 10.1% | 1,805 | 8.0% |
|  | 45-49 |  | 5,611 | 10.8% | 7,980 | 9.8% | 1714 | 8.7% | 2,172 | 9.6% |
|  | 50-54 |  | 6,153 | 11.8% | 11,059 | 13.5% | 1955 | 9.9% | 3,024 | 13.4% |
|  | 55-59 |  | 6,281 | 12.0% | 14,123 | 17.3% | 2035 | 10.3% | 3,833 | 17.0% |
|  | 60-64 |  | 5,739 | 11.0% | 16,482 | 20.1% | 1982 | 10.0% | 4,530 | 20.1% |
| **Ethnicity** | Missing |  | 3,714 | 7.1% | 3,938 | 4.8% | 1,074 | 5.4% | 1,182 | 5.2% |
|  | African |  | 1,597 | 3.1% | 1,529 | 1.9% | 606 | 3.1% | 378 | 1.7% |
|  | Any other Asian background |  | 1136 | 2.2% | 1,240 | 1.5% | 397 | 2.0% | 324 | 1.4% |
|  | Any other Black background |  | 869 | 1.7% | 735 | 0.9% | 224 | 1.1% | 175 | 0.8% |
|  | Any other White background |  | 5,296 | 10.2% | 5,160 | 6.3% | 1,845 | 9.3% | 1,407 | 6.2% |
|  | Any other ethnic group |  | 1,669 | 3.2% | 1,460 | 1.8% | 608 | 3.1% | 415 | 1.8% |
|  | Any other mixed background |  | 501 | 1.0% | 467 | 0.6% | 173 | 0.9% | 113 | 0.5% |
|  | Bangladeshi or British Bangladeshi |  | 731 | 1.4% | 636 | 0.8% | 250 | 1.3% | 147 | 0.7% |
|  | British, Mixed British |  | 30,215 | 57.9% | 60,081 | 73.4% | 12455 | 62.9% | 16661 | 73.9% |
|  | Caribbean |  | 1,356 | 2.6% | 857 | 1.0% | 289 | 1.5% | 206 | 0.9% |
|  | Chinese |  | 245 | 0.5% | 184 | 0.2% | 60 | 0.3% | 71 | 0.3% |
|  | Indian or British Indian |  | 1,522 | 2.9% | 2,245 | 2.7% | 646 | 3.3% | 616 | 2.7% |
|  | Irish |  | 208 | 0.4% | 590 | 0.7% | 117 | 0.6% | 157 | 0.7% |
|  | Pakistani or British Pakistani |  | 2,360 | 4.5% | 2,006 | 2.5% | 786 | 4.0% | 528 | 2.3% |
|  | White and Asian |  | 181 | 0.3% | 184 | 0.2% | 69 | 0.3% | 45 | 0.2% |
|  | White and Black African |  | 191 | 0.4% | 188 | 0.2% | 66 | 0.3% | 55 | 0.2% |
|  | White and Black Caribbean |  | 364 | 0.7% | 321 | 0.4% | 125 | 0.6% | 68 | 0.3% |
| **NHS Region** | East of England |  | 4,507 | 8.6% | 7,886 | 9.6% | 1,799 | 9.1% | 2,067 | 9.2% |
|  | London |  | 8,343 | 16.0% | 10,895 | 13.3% | 3,314 | 16.7% | 2,703 | 12.0% |
|  | Midlands |  | 11,370 | 21.8% | 17,289 | 21.1% | 4,160 | 21.0% | 5,060 | 22.4% |
|  | North East |  | 9,393 | 18.0% | 13,400 | 16.4% | 3,260 | 16.5% | 3,813 | 16.9% |
|  | North West |  | 8,423 | 16.1% | 13,772 | 16.8% | 3,104 | 15.7% | 3,669 | 16.3% |
|  | South East |  | 5,463 | 10.5% | 11,085 | 13.5% | 2609 | 13.2% | 3,146 | 14.0% |
|  | South West |  | 4,656 | 8.9% | 7,494 | 9.2% | 1544 | 7.8% | 2,090 | 9.3% |
| **IMD Quintiles** | 1 |  | 16,559 | 31.7% | 24,343 | 29.8% | 6,103 | 30.8% | 6,675 | 29.6% |
|  | 2 |  | 12,173 | 23.3% | 18,626 | 22.8% | 4,503 | 22.8% | 5,061 | 22.4% |
|  | 3 |  | 9,193 | 17.6% | 14,926 | 18.2% | 3,600 | 18.2% | 4,182 | 18.5% |
|  | 4 |  | 7,621 | 14.6% | 12,983 | 15.9% | 2,930 | 14.8% | 3,697 | 16.4% |
|  | 5 |  | 6,259 | 12.0% | 10,408 | 12.7% | 2,552 | 12.9% | 2,836 | 12.6% |
|  | Missing |  | 350 | 0.7% | 535 | 0.7% | 102 | 0.5% | 97 | 0.4% |
| **Previously positive** | No |  | 51,292 | 98.3% | 71,560 | 87.5% | 17,583 | 88.8% | 18,748 | 83.1% |
|  | Yes |  | 863 | 1.7% | 10,261 | 12.5% | 2,207 | 11.2% | 3,800 | 16.9% |
| **Vaccine priority groups** | Healthcare worker |  | 1,354 | 2.6% | 3,216 | 3.9% | 1,098 | 5.5% | 939 | 4.2% |
|  | CEV |  | 8,311 | 15.9% | 24,526 | 30.0% | 5,123 | 25.9% | 6,245 | 27.7% |
|  | At risk |  | 22,997 | 44.1% | 51,771 | 63.3% | 9,967 | 50.4% | 13,907 | 61.7% |
|  | Severely immunosuppressed |  | 1,458 | 2.8% | 4,939 | 6.0% | 1,550 | 7.8% | 1,352 | 6.0% |

### Table S2: Descriptive characteristics of positive and negative test results in individuals 65 years of age and over tested for SARS-CoV-2 in England for the study population

|  |  | |  | | **Delta** | | | | **Omicron** | | | |
| --- | --- | --- | --- | --- | --- | --- | --- | --- | --- | --- | --- | --- |
|  |  | |  | | **Positive** | | **Negative** | | **Positive** | | **Negative** | |
|  |  |  | |  | **n** | **%** | **n** | **%** | **n** | **%** | **n** | **%% %** |
| **Category** |  | |  | | **30,427** |  | **187,463** |  | **13,348** |  | **49,643** |  |
| **Vaccination Status after vaccine** | **Manufacturer (primary course)** | | **Dose** | |  |  |  |  |  |  |  |  |
|  | Unvaccinated | | | | 5,033 | 16.5% | 6676 | 3.3% | 1452 | 1.1% | 1462 | 3% |
|  | ChAdOx1-S | | Dose 1 | | 590 | 1.9% | 5,399 | 2.7% | 153 | 9.7% | 331 | 1% |
|  |  | | Dose 2 | | 14073 | 46.3% | 70923 | 35.0% | 1295 | 35.1% | 4057 | 8% |
|  |  | | Booster (any) | | 921 | 3.0% | 19693 | 9.7% | 4679 | 1.0% | 20015 | 40% |
|  | BNT162b2 | | Dose 1 | | 336 | 1.1% | 2237 | 1.1% | 133 | 4.5% | 256 | 1% |
|  |  | | Dose 2 | | 8270 | 27.2% | 58131 | 28.7% | 607 | 37.6% | 2275 | 5% |
|  |  | | Booster (any) | | 1197 | 3.9% | 24358 | 12.0% | 5016 | 0.1% | 21227 | 43% |
|  | mRNA-1273 | | Dose 1 | | 6 | 0.0% | 30 | 0.0% | 11 | 0.0% | 11 | 0% |
|  |  | | Dose 2 | | 1 | 0.0% | 15 | 0.0% | 2 | 0.0% | 8 | 0% |
|  |  | | Booster (any) | | 0 | 0.0% | 1 | 0.0% | 0 | 8.8% | 1 | 0% |
| **Outcomes** | ECDS admissions -pillar 2 symptomatic | | | | 5,331 | 100.0% | 6,059 | 100.0% | 1174 | 100.0% | 2,317 | 100.0% |
|  | ECDS admissions -pillar 2 symptomatic with respiratory code | | | | 3,170 | 59.5% | 1,559 | 25.7% | 467 | 39.8% | 547 | 23.6% |
|  | Secondary Uses Service Respiratory coded admissions (SUS) | | | | 29,596 | 100.0% | 183,108 | 100.0% | 12,250 | 100.0% | 47,931 | 100.0% |
|  | Secondary Uses Service Respiratory coded admissions (SUS) with 0 days length of stay | | | | 989 | 3.3% | 6,450 | 3.5% | 1,393 | 11.4% | 2,121 | 4.4% |
|  | SUS with respiratory code but NOT in primary diagnosis field with at least 1 day stay | | | | 5,446 | 18.4% | 82,963 | 45.3% | 4,704 | 38.4% | 20,067 | 41.9% |
|  | SUS with respiratory code as the primary diagnosis and length of stay >=1 day | | | | 23,161 | 78.3% | 93,695 | 51.2% | 6,153 | 50.2% | 25,743 | 53.7% |
|  | SUS with respiratory code as the primary diagnosis and length of stay >=2 days | | | | 21,808 | 73.7% | 85,806 | 46.9% | 5,565 | 45.4% | 23,257 | 48.5% |
|  | SUS with respiratory code as the primary diagnosis and length of stay >=3 days | | | | 20,271 | 68.5% | 77,998 | 42.6% | 5,019 | 41.0% | 20,796 | 43.4% |
|  | SUS >=2 days stay and primary diagnosis with oxygen use | | | | 3,625 | 12.2% | 4,092 | 2.2% | 517 | 4.2% | 1,155 | 2.4% |
|  | SUS >=2 days stay and primary diagnosis with oxygen use or ventilation used or in ICU | | | | 6,265 | 21.2% | 7,146 | 3.9% | 807 | 6.6% | 1,921 | 4.0% |
| **Testing route** | Pillar 1 | |  | | 20,659 | 67.9% | 179,756 | 95.9% | 9,482 | 71.0% | 46,858 | 94.4% |
|  | Pillar 2 | |  | | 9,768 | 32.1% | 7,707 | 4.1% | 3,866 | 29.0% | 2,785 | 5.6% |
| **Gender** | Female | |  | | 13,739 | 45.2% | 94,863 | 50.6% | 6,379 | 47.8% | 25,660 | 51.7% |
|  | Male | |  | | 16,668 | 54.8% | 91,485 | 48.8% | 6,947 | 52.0% | 23,756 | 47.9% |
|  | Missing | |  | | 20 | 0.1% | 1,115 | 0.6% | 22 | 0.2% | 227 | 0.5% |
| **Age** | 65-69 | |  | | 5,515 | 18.1% | 19,569 | 10.4% | 1,932 | 14.5% | 5,375 | 10.8% |
|  | 70-74 | |  | | 6,456 | 21.2% | 28,250 | 15.1% | 2,368 | 17.7% | 7,556 | 15.2% |
|  | 75-79 | |  | | 6,021 | 19.8% | 33,227 | 17.7% | 2,540 | 19.0% | 8,899 | 17.9% |
|  | 80-84 | |  | | 5,296 | 17.4% | 36,800 | 19.6% | 2,534 | 19.0% | 9,906 | 20.0% |
|  | 85-89 | |  | | 4,255 | 14.0% | 36,298 | 19.4% | 2,251 | 16.9% | 9,571 | 19.3% |
|  | >=90 | |  | | 2,884 | 9.5% | 33,319 | 17.8% | 1,723 | 12.9% | 8,336 | 16.8% |
| **Ethnicity** | Missing | |  | | 1,412 | 4.6% | 7,838 | 4.2% | 579 | 4.3% | 2,221 | 4.5% |
|  | African | |  | | 238 | 0.8% | 597 | 0.3% | 105 | 0.8% | 131 | 0.3% |
|  | Any other Asian background | |  | | 308 | 1.0% | 1,165 | 0.6% | 134 | 1.0% | 294 | 0.6% |
|  | Any other Black background | |  | | 152 | 0.5% | 298 | 0.2% | 50 | 0.4% | 79 | 0.2% |
|  | Any other White background | |  | | 1,559 | 5.1% | 8,230 | 4.4% | 684 | 5.1% | 2,163 | 4.4% |
|  | Any other ethnic group | |  | | 354 | 1.2% | 1,297 | 0.7% | 145 | 1.1% | 314 | 0.6% |
|  | Any other mixed background | |  | | 79 | 0.3% | 469 | 0.3% | 41 | 0.3% | 102 | 0.2% |
|  | Bangladeshi or British Bangladeshi | |  | | 207 | 0.7% | 518 | 0.3% | 68 | 0.5% | 114 | 0.2% |
|  | British, Mixed British | |  | | 23,386 | 76.9% | 158,305 | 84.4% | 10,445 | 78.3% | 42,004 | 84.6% |
|  | Caribbean | |  | | 645 | 2.1% | 1,179 | 0.6% | 248 | 1.9% | 273 | 0.5% |
|  | Chinese | |  | | 92 | 0.3% | 244 | 0.1% | 32 | 0.2% | 57 | 0.1% |
|  | Indian or British Indian | |  | | 797 | 2.6% | 2,941 | 1.6% | 292 | 2.2% | 798 | 1.6% |
|  | Irish | |  | | 317 | 1.0% | 2,370 | 1.3% | 186 | 1.4% | 585 | 1.2% |
|  | Pakistani or British Pakistani | |  | | 740 | 2.4% | 1,671 | 0.9% | 296 | 2.2% | 428 | 0.9% |
|  | White and Asian | |  | | 37 | 0.1% | 109 | 0.1% | 9 | 0.1% | 26 | 0.1% |
|  | White and Black African | |  | | 24 | 0.1% | 79 | 0.0% | 5 | 0.0% | 20 | 0.0% |
|  | White and Black Caribbean | |  | | 80 | 0.3% | 153 | 0.1% | 29 | 0.2% | 34 | 0.1% |
| **-NHS Region** | East of England | |  | | 2,666 | 8.8% | 20,728 | 11.1% | 1,227 | 9.2% | 5,013 | 10.1% |
|  | London | |  | | 3,670 | 12.1% | 20,657 | 11.0% | 1,952 | 14.6% | 4,858 | 9.8% |
|  | Midlands | |  | | 6,224 | 20.5% | 37,399 | 20.0% | 2,643 | 19.8% | 11,046 | 22.3% |
|  | North East | |  | | 6,832 | 22.5% | 29,448 | 15.7% | 2,501 | 18.7% | 8,136 | 16.4% |
|  | North West | |  | | 4,838 | 15.9% | 28,624 | 15.3% | 2,180 | 16.3% | 7,386 | 14.9% |
|  | South East | |  | | 3,305 | 10.9% | 30,103 | 16.1% | 1,859 | 13.9% | 7,909 | 15.9% |
|  | South West | |  | | 2,892 | 9.5% | 20,504 | 10.9% | 986 | 7.4% | 5,295 | 10.7% |
| **IMD Quintiles** | 1 | |  | | 8,146 | 26.8% | 37,552 | 20.0% | 3,379 | 25.3% | 9,855 | 19.9% |
|  | 2 | |  | | 6,581 | 21.6% | 37,515 | 20.0% | 2,726 | 20.4% | 9,933 | 20.0% |
|  | 3 | |  | | 5,734 | 18.8% | 38,002 | 20.3% | 2,626 | 19.7% | 10,023 | 20.2% |
|  | 4 | |  | | 5,457 | 17.9% | 38,607 | 20.6% | 2,423 | 18.2% | 10,333 | 20.8% |
|  | 5 | |  | | 4,441 | 14.6% | 35,434 | 18.9% | 2,165 | 16.2% | 9,420 | 19.0% |
|  | Missing | |  | | 68 | 0.2% | 353 | 0.2% | 29 | 0.2% | 79 | 0.2% |
| **Previously positive** | No | |  | | 29,963 | 41.6% | 171,099 | 23.9% | 12,649 | 94.8% | 44,761 | 90.2% |
|  | Yes | |  | | 464 | 2.3% | 16,364 | 2.6% | 699 | 5.2% | 4,882 | 9.8% |
| **Vaccine priority groups** | Healthcare worker | |  | | 63 | 0.2% | 262 | 0.1% | 30 | 0.2% | 74 | 0.1% |
|  | CEV | |  | | 13,802 | 45.4% | 98,148 | 52.4% | 7,054 | 52.8% | 24,200 | 48.7% |
|  | Severely immunosuppressed | |  | | 1,893 | 6.2% | 11,918 | 6.4% | 1,434 | 10.7% | 3,598 | 7.2% |

### Table S3: Vaccine effectiveness (and 95% CI) at least 7 days after the third dose of COVID-19 vaccine using ECDS and secondary care hospital admission data by the Delta and Omicron variants by age group

| **Age 18-64** | **Omicron** | | | **Delta** | | |
| --- | --- | --- | --- | --- | --- | --- |
| **Outcome** | **cases** | **controls** | **VE ≥7 days post booster** | **cases** | **controls** | **VE ≥7 days post booster** |
| **P2s all cases and controls** | 713,260 | 827,973 | 61.9 (61.5 to 62.3) | 18,686 | 560,146 | 94.2 (94.1 to 94.3) |
| **P2s ECDS (all controls)** | 1,472 | 827,973 | 80.0 (77.8 to 82.0) | 115 | 560,146 | 99.1 (98.9 to 99.3) |
| **P2s ECDS (ECDS controls)** | 1,472 | 3,268 | 75.9 (71.7 to 79.5) | 115 | 2,167 | 98.7 (98.4 to 99.0) |
| **P2s ECDS ARI coded (all controls)** | 323 | 827,973 | 86.9 (83.8 to 89.4) | 54 | 560,146 | 99.4 (99.2 to 99.5) |
| **P2s ECDS ARI coded (ECDS ARI controls)** | 323 | 479 | 87.4 (80.8 to 91.8) | 54 | 361 | 99.3 (99.0 to 99.5) |
| **P1/2 SUS 0 days** | 3,291 | 1,061 | 63.9 (55.0 to 71.1) | 36 | 740 | 93.6 (90.6 to 95.7) |
| **P1/2 SUS 1+ days Not Primary** | 2,201 | 3,408 | 55.2 (48.9 to 60.7) | 63 | 2,425 | 87.3 (83.0 to 90.5) |
| **P1/2 SUS 1+ days Primary** | 1,436 | 3,462 | 82.3 (79.3 to 84.9) | 192 | 2,658 | 96.4 (95.6 to 97.0) |
| **P1/2 SUS 2+ days Primary** | 921 | 2,834 | 84.5 (81.5 to 87.1) | 173 | 2,209 | 96.3 (95.5 to 97.0) |
| **P1/2 SUS 3+ days Primary** | 640 | 2,337 | 86.1 (83.0 to 88.6) | 154 | 1,859 | 96.4 (95.5 to 97.1) |
| **P1/2 SUS 2+ days Primary & oxygen** | 46 | 168 | 93.9 (85.0 to 97.5) | 42 | 133 | 94.4 (89.8 to 96.9) |
| **P1/2 SUS 2+ days Primary & oxygen, ventilation, or ICU** | 87 | 368 | 94.0 (89.3 to 96.6) | 67 | 297 | 93.9 (90.7 to 96.1) |
| **Age 65+** | **Omicron** | | | **Delta** | | |
| **Outcome** | **cases** | **controls** | **VE ≥7 days post booster** | **cases** | **controls** | **VE ≥7 days post booster** |
| **P2s all cases and controls** | 102,720 | 137,812 | 52.3 (48.3 to 55.9) | 7,384 | 130,830 | 91.1 (90.6 to 91.6) |
| **P2s ECDS (all controls)** | 908 | 137,812 | 91.3 (88.9 to 93.1) | 203 | 130,830 | 98.8 (98.6 to 99) |
| **P2s ECDS (ECDS controls)** | 908 | 2,034 | 85.7 (77.5 to 90.9) | 203 | 1,941 | 98.4 (97.8 to 98.8) |
| **P2s ECDS ARI coded (all controls)** | 358 | 137,812 | 93.4 (90.6 to 95.3) | 105 | 130,830 | 99.1 (98.8 to 99.3) |
| **P2s ECDS ARI coded (ECDS ARI controls)** | 358 | 481 | 95.2 (87.0 to 98.2) | 105 | 521 | 98.6 (97.7 to 99.2) |
| **P1/2 SUS 0 days** | 1,203 | 1,778 | 71.6 (54.4 to 82.3) | 50 | 1,568 | 88.4 (81.6 to 92.6) |
| **P1/2 SUS 1+ days Not Primary** | 3,595 | 16,111 | 62.0 (54.8 to 68) | 318 | 15,966 | 81.4 (77.7 to 84.4) |
| **P1/2 SUS 1+ days Primary** | 3,934 | 20,822 | 87.7 (85.9 to 89.3) | 981 | 21,215 | 94.5 (93.9 to 95.0) |
| **P1/2 SUS 2+ days Primary** | 3,501 | 18,765 | 88.3 (86.4 to 89.9) | 913 | 19,259 | 94.5 (93.8 to 95.0) |
| **P1/2 SUS 3+ days Primary** | 3,109 | 16,702 | 88.7 (86.8 to 90.4) | 847 | 17,266 | 94.3 (93.6 to 94.9) |
| **P1/2 SUS 2+ days Primary & oxygen** | 287 | 913 | 93.3 (88.2 to 96.2) | 145 | 997 | 96.2 (94.6 to 97.4) |
| **P1/2 SUS 2+ days Primary & oxygen, ventilation, or ICU** | 451 | 1,516 | 92.5 (88.5 to 95.1) | 247 | 1,642 | 95.9 (94.7 to 96.9) |

P2s: Pillar 2 symptomatic	P1/2: Pillar 1 and 2	SUS: Secondary User Service	ECDS: Emergency care data set

Primary: respiratory discharge code in the primary field	Not Primary: respiratory discharge code not in the primary field

0 days, 1+ days, 2+ days, 3+ days: Length of stay, for example 1+ means at least one overnight stay

### Table S4: Vaccine effectiveness (95% CI) against different hospitalisation outcomes with Delta by dose and interval (all vaccines combined)

|  |  | P2s ECDS (ECDS controls) | P2s ECDS ARI coded (ECDS ARI controls) | P1/2 SUS 0 days | P1/2 SUS 1+ days Not Primary | P1/2 SUS 1+ days Primary | P1/2 SUS 2+ days Primary | P1/2 SUS 3+ days Primary | P1/2 SUS 2+ days Primary & oxygen | P1/2 SUS 2+ days Primary & oxygen, ventilation, or ICU |
| --- | --- | --- | --- | --- | --- | --- | --- | --- | --- | --- |
|  | **18-64** | | | | | | | | | |
| Dose 1 | 0-27 | 67.1 (61.8 to 71.6) | 69.6 (58.4 to 77.7) | 35.8 (14.8 to 51.6) | 40.3 (24.8 to 52.6) | 44.7 (34.3 to 53.5) | 43.5 (31.1 to 53.6) | 46.3 (32.8 to 57.1) | 41.4 (-14.8 to 70.1) | 51.0 (18.7 to 70.4) |
|  | 28+ | 85 (83.4 to 86.5) | 89.2 (86.6 to 91.4) | 65.1 (59.0 to 70.3) | 54.7 (49.5 to 59.3) | 82.7 (81.0 to 84.3) | 82.7 (80.8 to 84.4) | 83.0 (80.9 to 84.8) | 83.3 (76.7 to 88.0) | 82.7 (78.0 to 86.4) |
| Dose 2 | 0-13 | 92.8 (91.3 to 94.1) | 94.6 (92.0 to 96.4) | 83.4 (76.9 to 88.1) | 63.2 (51.6 to 72.0) | 90.5 (88.1 to 92.5) | 90.8 (88.0 to 92.9) | 91.1 (88.2 to 93.3) | 80.0 (55.7 to 91.0) | 85.0 (73.0 to 91.7) |
|  | 14-174 | 93.1 (92.5 to 93.6) | 95.0 (94.1 to 95.8) | 82.5 (80.3 to 84.4) | 71.1 (68.7 to 73.2) | 91.0 (90.4 to 91.5) | 91.1 (90.5 to 91.7) | 91.0 (90.3 to 91.7) | 91.4 (89.2 to 93.1) | 90.5 (88.9 to 91.9) |
|  | 175+ | 90.4 (89.2 to 91.5) | 93.3 (91.5 to 94.7) | 75.0 (69.7 to 79.3) | 61.8 (56.1 to 66.7) | 84.2 (82.5 to 85.7) | 84.5 (82.7 to 86.1) | 85.2 (83.3 to 86.8) | 86.1 (80.3 to 90.2) | 84.3 (79.9 to 87.7) |
| Booster | 0-6 | 94.7 (93.3 to 95.9) | 97.0 (95.4 to 98.0) | 86.7 (78.2 to 91.8) | 76.7 (64.6 to 84.7) | 88.9 (85.7 to 91.5) | 90.0 (86.7 to 92.5) | 90.3 (86.8 to 92.9) | 94.0 (83.9 to 97.8) | 92.7 (86.1 to 96.1) |
|  | 7-13 | 98.5 (97.7 to 99.0) | 99.3 (98.6 to 99.6) | 92.5 (83.9 to 96.5) | 86.4 (76.9 to 92.1) | 95.0 (93.1 to 96.4) | 94.8 (92.6 to 96.3) | 94.7 (92.4 to 96.4) | 95.0 (81.4 to 98.6) | 93.9 (87.2 to 97.1) |
|  | 14-34 | 99.0 (98.6 to 99.3) | 99.5 (99.1 to 99.7) | 95.3 (91.4 to 97.4) | 87.3 (80.9 to 91.6) | 97.5 (96.7 to 98.2) | 97.4 (96.4 to 98.1) | 97.6 (96.6 to 98.3) | 96.0 (90.4 to 98.3) | 95.2 (90.7 to 97.5) |
|  | 35-69 | 98.5 (97.9 to 99.0) | 99.0 (98.1 to 99.4) | 94.0 (88.0 to 97.0) | 87.4 (77.4 to 93.0) | 96.0 (94.3 to 97.2) | 96.1 (94.3 to 97.3) | 96.0 (94.1 to 97.4) | 93.2 (83.4 to 97.3) | 93.5 (86.2 to 97.0) |
|  | 70-104 | 98.7 (95.9 to 99.6) | 99.1 (95.5 to 99.8) | 71.8 (21.2 to 89.9) |  | 89.7 (75.9 to 95.6) | 91.3 (75.4 to 96.9) | 91.7 (76.0 to 97.1) |  |  |
|  | **65+** | | | | | | | | | |
| Dose 1 | 0-27 |  |  |  |  | 55.1 (35.7 to 68.7) | 59.2 (41.1 to 71.8) | 55.0 (33.8 to 69.4) |  |  |
|  | 28+ | 84.5 (76.1 to 90) | 89.8 (79.1 to 95) | 65.4 (37.0 to 81.0) | 40.3 (27.4 to 50.9) | 74.7 (71.6 to 77.6) | 74.5 (71.2 to 77.4) | 74.2 (70.7 to 77.3) | 82.3 (71.7 to 89.0) | 86.7 (81.4 to 90.5) |
| Dose 2 | 0-13 | 85.1 (60.7 to 94.3) | 81.4 (26.7 to 95.3) | 66.8 (-64.5 to 93.3) | 48.3 (6.8 to 71.4) | 82.8 (75.0 to 88.2) | 81.8 (73.3 to 87.6) | 82.7 (74.0 to 88.5) | 88.1 (64.4 to 96.0) | 89.8 (74.5 to 95.9) |
|  | 14-174 | 89.6 (86.6 to 91.9) | 89.1 (83.0 to 93.0) | 76.0 (67.0 to 82.6) | 57.7 (52.4 to 62.4) | 85.7 (84.7 to 86.6) | 85.6 (84.6 to 86.6) | 85.7 (84.7 to 86.7) | 90.1 (87.3 to 92.2) | 90.5 (88.6 to 92.1) |
|  | 175+ | 85.0 (80.5 to 88.5) | 85.1 (76.3 to 90.6) | 56.6 (38.8 to 69.2) | 46.7 (39.3 to 53.1) | 78.2 (76.6 to 79.8) | 78.3 (76.6 to 79.9) | 78.1 (76.3 to 79.7) | 83.9 (78.8 to 87.8) | 84.9 (81.5 to 87.7) |
| Booster | 0-6 | 91.7 (88.3 to 94.1) | 91.9 (85.3 to 95.5) | 81.8 (66.9 to 90.0) | 54.4 (43.1 to 63.5) | 86.8 (84.8 to 88.5) | 86.9 (84.8 to 88.6) | 86.4 (84.2 to 88.3) | 93.5 (89.2 to 96.0) | 92.1 (88.4 to 94.6) |
|  | 7-13 | 97.3 (96.0 to 98.2) | 97.6 (95.3 to 98.8) | 77.8 (58.6 to 88.1) | 71.3 (63.2 to 77.7) | 88 (86.2 to 89.6) | 88.6 (86.8 to 90.1) | 88.3 (86.4 to 90.0) | 91.6 (86.1 to 94.9) | 92.3 (88.9 to 94.7) |
|  | 14-34 | 98.9 (98.4 to 99.3) | 99.0 (98.2 to 99.4) | 90.8 (83.7 to 94.8) | 83.9 (80.0 to 87.1) | 96.4 (95.8 to 96.9) | 96.3 (95.7 to 96.8) | 96.1 (95.5 to 96.6) | 97.7 (96.3 to 98.5) | 97.5 (96.5 to 98.3) |
|  | 35-69 | 98.4 (97.7 to 98.9) | 98.7 (97.5 to 99.3) | 90.9 (81.1 to 95.6) | 85.4 (80.5 to 89.1) | 95.7 (94.9 to 96.4) | 95.7 (94.8 to 96.4) | 95.6 (94.7 to 96.3) | 96.7 (94.4 to 98.1) | 96.1 (94.0 to 97.4) |
|  | 70-104 | 97.7 (95.5 to 98.8) | 98.0 (94.6 to 99.3) |  | 55.7 (-23.9 to 84.2) | 91.2 (85.2 to 94.8) | 90.2 (83.5 to 94.3) | 89.7 (82.1 to 94.1) | 90.0 (43.4 to 98.2) | 87.0 (58.6 to 95.9) |

P2s: Pillar 2 symptomatic	P1/2: Pillar 1 and 2	SUS: Secondary User Service	ECDS: Emergency care data set

Primary: respiratory discharge code in the primary field	Not Primary: respiratory discharge code not in the primary field

0 days, 1+ days, 2+ days, 3+ days: Length of stay, for example 1+ means at least one overnight stay

Table S5: Vaccine effectiveness against hospital admissions from emergency care (ECDS) within 14 days of the test date by the Delta and Omicron variants in symptomatic individuals 18 to 64 years of age

|  | **Omicron** | | | | | | |
| --- | --- | --- | --- | --- | --- | --- | --- |
|  |  | **Pillar 2 symptomatic with ECDS admission within 14 days** | | | **Pillar 2 symptomatic with ARI coded ECDS admission within 14 days** | | |
|  | **Interval (days)** | **cases** | **controls** | **VE (95% CI)** | **cases** | **controls** | **VE (95% CI)** |
| **unvaccinated** | | **653** | **497** |  | **177** | **66** |  |
| Dose 1 | 0-27 | 28 | 43 | 48.5 (12.3 to 69.7) | 6 | 4 |  |
|  | 28+ | 148 | 226 | 48.7 (32.8 to 60.8) | 28 | 39 | 75.0 (50.3 to 87.4) |
| Dose 2 | 0-13 | 13 | 24 | 39.6 (-31.5 to 72.2) | 3 | 5 |  |
|  | 14-174 | 441 | 1,111 | 54.7 (45.3 to 62.4) | 75 | 188 | 73.7 (56.9 to 84.0) |
|  | 175+ | 659 | 968 | 34.6 (21.7 to 45.4) | 152 | 142 | 46.5 (14.2 to 66.7) |
| Booster | 0-6 | 105 | 295 | 63.9 (52.2 to 72.8) | 18 | 50 | 76.3 (50.5 to 88.7) |
|  | 7-13 | 88 | 328 | 80.1 (73.5 to 85.1) | 17 | 67 | 91.4 (82.7 to 95.7) |
|  | 14-34 | 288 | 1,013 | 82.4 (78.6 to 85.6) | 52 | 148 | 91.4 (85.5 to 94.9) |
|  | 35-69 | 584 | 1,248 | 72.7 (67.2 to 77.2) | 116 | 178 | 86.2 (77.8 to 91.5) |
|  | 70-104 | 377 | 538 | 66.9 (59.1 to 73.3) | 100 | 69 | 79.5 (64.5 to 88.1) |
|  | 105+ | 135 | 141 | 53.6 (36.9 to 65.9) | 38 | 17 | 60.7 (14.7 to 81.9) |
|  | **Delta** | | | | | | |
|  | **Interval (days)** | **cases** | **controls** | **VE (95% CI)** | **cases** | **controls** | **VE (95% CI)** |
| **unvaccinated** | | **9,170** | **2,696** |  | **5,447** | **381** |  |
| Dose 1 | 0-27 | 486 | 574 | 67.1 (61.8 to 71.6) | 262 | 81 | 69.6 (58.4 to 77.7) |
|  | 28+ | 999 | 1,868 | 85.0 (83.4 to 86.5) | 456 | 279 | 89.2 (86.6 to 91.4) |
| Dose 2 | 0-13 | 161 | 551 | 92.8 (91.3 to 94.1) | 76 | 78 | 94.6 (92.0 to 96.4) |
|  | 14-174 | 4,982 | 9,434 | 93.1 (92.5 to 93.6) | 2,611 | 1,511 | 95.0 (94.1 to 95.8) |
|  | 175+ | 1,192 | 1,786 | 90.4 (89.2 to 91.5) | 643 | 304 | 93.3 (91.5 to 94.7) |
| Booster | 0-6 | 127 | 385 | 94.7 (93.3 to 95.9) | 69 | 75 | 97.0 (95.4 to 98.0) |
|  | 7-13 | 32 | 394 | 98.5 (97.7 to 99.0) | 13 | 85 | 99.3 (98.6 to 99.6) |
|  | 14-34 | 43 | 897 | 99.0 (98.6 to 99.3) | 19 | 144 | 99.5 (99.1 to 99.7) |
|  | 35-69 | 37 | 694 | 98.5 (97.9 to 99.0) | 20 | 104 | 99.0 (98.1 to 99.4) |
|  | 70-104 | 3 | 179 | 98.7 (95.9 to 99.6) | 2 | 26 | 99.1 (95.5 to 99.8) |
|  | 105+ | 0 | 3 |  | 0 | 2 |  |

### Table S6: Vaccine effectiveness against hospital admissions from emergency care (ECDS) within 14 days of the test date by the Delta and Omicron variants in symptomatic individuals 18 to 64 years of age using all controls

|  | **Omicron** | | | | | | |
| --- | --- | --- | --- | --- | --- | --- | --- |
|  |  | **Pillar 2 symptomatic with ECDS admission within 14 days** | | | **Pillar 2 symptomatic with ARI coded ECDS admission within 14 days** | | |
|  | **Interval (days)** | **cases** | **controls** | **VE (95% CI)** | **cases** | **controls** | **VE (95% CI)** |
| **unvaccinated** | | **653** | **127,778** |  | **177** | **127,778** |  |
| Dose 1 | 0-27 | 28 | 9832 | 41.1 (13.8 to 59.7) | 6 | 9,832 | 50.6 (-11.9 to 78.2) |
|  | 28+ | 148 | 48,886 | 39.2 (27.0 to 49.3) | 28 | 48,886 | 58.6 (37.6 to 72.5) |
| Dose 2 | 0-13 | 13 | 7354 | 56.9 (25.2 to 75.2) | 3 | 7,354 | 60.9 (-22.7 to 87.6) |
|  | 14-174 | 441 | 440,896 | 59.3 (53.9 to 64.1) | 75 | 440,896 | 71.7 (62.5 to 78.6) |
|  | 175+ | 659 | 212,640 | 33.4 (25.3 to 40.6) | 152 | 212,640 | 50.3 (37.5 to 60.5) |
| Booster | 0-6 | 105 | 100,217 | 67.8 (60.2 to 73.9) | 18 | 100,217 | 80.1 (67.3 to 87.9) |
|  | 7-13 | 88 | 106,474 | 83.3 (79.0 to 86.7) | 17 | 106,474 | 88.9 (81.4 to 93.4) |
|  | 14-34 | 288 | 281,633 | 85.5 (83.3 to 87.5) | 52 | 281,633 | 91.1 (87.7 to 93.6) |
|  | 35-69 | 584 | 306,848 | 79.1 (76.3 to 81.5) | 116 | 306,848 | 87.7 (84.1 to 90.5) |
|  | 70-104 | 377 | 106,966 | 70.6 (66.0 to 74.6) | 100 | 106,966 | 79.5 (72.8 to 84.6) |
|  | 105+ | 135 | 26,052 | 60.5 (51.2 to 68.1) | 38 | 26,052 | 72.5 (58.6 to 81.8) |
|  | **Delta** | | | | | | |
|  | **Interval (days)** | **cases** | **controls** | **VE (95% CI)** | **cases** | **controls** | **VE (95% CI)** |
| **unvaccinated** | | **9170** | **566,636** |  | **5447** | **566,636** |  |
| Dose 1 | 0-27 | 486 | 159,118 | 74.1 (71.6 to 76.5) | 262 | 159,118 | 75.3 (71.9 to 78.3) |
|  | 28+ | 999 | 391,467 | 86.9 (86.0 to 87.8) | 456 | 391,467 | 89.9 (88.9 to 90.9) |
| Dose 2 | 0-13 | 161 | 122309 | 94.2 (93.2 to 95.0) | 76 | 122,309 | 95.6 (94.4 to 96.5) |
|  | 14-174 | 4982 | 2,218,985 | 94.7 (94.5 to 94.9) | 2611 | 2,218,985 | 95.7 (95.4 to 95.9) |
|  | 175+ | 1192 | 363,447 | 92.0 (91.4 to 92.5) | 643 | 363,447 | 93.1 (92.5 to 93.7) |
| Booster | 0-6 | 127 | 117,633 | 95.9 (95.1 to 96.6) | 69 | 117,633 | 96.5 (95.6 to 97.3) |
|  | 7-13 | 32 | 117,498 | 98.9 (98.5 to 99.2) | 13 | 117,498 | 99.3 (98.8 to 99.6) |
|  | 14-34 | 43 | 236,845 | 99.3 (99.1 to 99.5) | 19 | 236,845 | 99.5 (99.2 to 99.7) |
|  | 35-69 | 37 | 157,782 | 99.0 (98.6 to 99.3) | 20 | 157,782 | 99.1 (98.7 to 99.5) |
|  | 70-104 | 3 | 47,688 | 99.3 (97.8 to 99.8) | 2 | 47,688 | 99.2 (97.0 to 99.8) |
|  | 105+ | 0 | 333 |  | 0 | 333 |  |

### Table S7: Vaccine effectiveness against hospital admissions from emergency care (ECDS) within 14 days of the test date by the Delta and Omicron variants in symptomatic individuals 65 years of age and older

| **Omicron** | | | | | | | |
| --- | --- | --- | --- | --- | --- | --- | --- |
|  |  | **Pillar 2 symptomatic with ECDS admission within 14 days** | | | **Pillar 2 symptomatic with ARI coded ECDS admission within 14 days** | | |
|  | **Interval (days)** | **cases** | **controls** | **VE (95% CI)** | **cases** | **controls** | **VE (95% CI)** |
| **unvaccinated** | | **104** | **39** |  | **49** | **10** |  |
| Dose 1 | 0-27 | 4 | 6 |  | 3 | 0 |  |
|  | 28+ | 17 | 13 |  | 6 | 2 |  |
| Dose 2 | 14-174 | 13 | 25 | 77.8 (45.0 to 91.0) | 5 | 8 | 90.6 (46.8 to 98.4) |
|  | 175+ | 119 | 160 | 66.7 (43.4 to 80.4) | 44 | 36 | 79.7 (34.4 to 93.7) |
| Booster | 0-6 | 9 | 40 | 85.8 (61.5 to 94.7) | 2 | 10 | 97.3 (79.2 to 99.7) |
|  | 7-13 | 6 | 56 | 92.3 (76.3 to 97.5) | 2 | 10 | 94.8 (-7.5 to 99.8) |
|  | 14-34 | 31 | 300 | 92.4 (86.0 to 95.8) | 10 | 86 | 97.9 (92.6 to 99.4) |
|  | 35-69 | 263 | 888 | 87.0 (79.2 to 91.8) | 101 | 218 | 95.3 (87.3 to 98.3) |
|  | 70-104 | 423 | 633 | 84.0 (74.6 to 89.9) | 166 | 137 | 94.2 (84.0 to 97.9) |
|  | 105+ | 185 | 157 | 76.9 (60.6 to 86.4) | 79 | 30 | 90.3 (67.8 to 97.1) |
| **Delta** | | | | | | | |
|  | **Interval (days)** | **cases** | **controls** | **VE (95% CI)** | **cases** | **controls** | **VE (95% CI)** |
| **unvaccinated** | | **701** | **115** |  | **458** | **32** |  |
| Dose 1 | 0-27 | 15 | 8 | 59.2 (-12.0 to 85.1) | 8 | 0 |  |
|  | 28+ | 78 | 130 | 84.5 (76.1 to 90.0) | 40 | 37 | 89.8 (79.1 to 95.0) |
| Dose 2 | 0-13 | 7 | 82 | 85.1 (60.7 to 94.3) | 5 | 17 | 81.4 (26.7 to 95.3) |
|  | 14-174 | 2,742 | 2,831 | 89.6 (86.6 to 91.9) | 1,634 | 690 | 89.1 (83.0 to 93.0) |
|  | 175+ | 1,417 | 780 | 85.0 (80.5 to 88.5) | 826 | 216 | 85.1 (76.3 to 90.6) |
| Booster | 0-6 | 168 | 172 | 91.7 (88.3 to 94.1) | 94 | 46 | 91.9 (85.3 to 95.5) |
|  | 7-13 | 57 | 172 | 97.3 (96.0 to 98.2) | 29 | 43 | 97.6 (95.3 to 98.8) |
|  | 14-34 | 63 | 583 | 98.9 (98.4 to 99.3) | 38 | 171 | 99.0 (98.2 to 99.4) |
|  | 35-69 | 68 | 864 | 98.4 (97.7 to 98.9) | 31 | 224 | 98.7 (97.5 to 99.3) |
|  | 70-104 | 15 | 322 | 97.7 (95.5 to 98.8) | 7 | 83 | 98.0 (94.6 to 99.3) |

### Table S8: Vaccine effectiveness against hospital admissions from emergency care (ECDS) within 14 days of the test date by the Delta and Omicron variants in symptomatic individuals 18 to 64 years of age using all controls

|  |  | **Pillar 2 symptomatic with ECDS admission within 14 days** | | | **Pillar 2 symptomatic with ARI coded ECDS admission within 14 days** | | |
| --- | --- | --- | --- | --- | --- | --- | --- |
|  | **Interval (days)** | **cases** | **controls** | **VE (95% CI)** | **cases** | **controls** | **VE (95% CI)** |
| **unvaccinated** | | **104** | **1,703** |  | **49** | **1,703** |  |
| Dose 1 | 0-27 | 4 | 211 | 58.4 (-23.1 to 85.9) | 3 | 211 | 16.7 (-182.9 to 75.5) |
|  | 28+ | 17 | 640 | 62.9 (35.5 to 78.7) | 6 | 640 | 73.8 (36.0 to 89.2) |
| Dose 2 | 0-13 | 0 | 30 |  | 0 | 30 |  |
|  | 14-174 | 13 | 977 | 80.8 (64.1 to 89.7) | 5 | 977 | 86.7 (64.6 to 95.0) |
|  | 175+ | 119 | 6,250 | 63.4 (50.5 to 72.9) | 44 | 6,250 | 76.6 (63.0 to 85.2) |
| Booster | 0-6 | 9 | 2,117 | 81.6 (62.0 to 91.1) | 2 | 2,117 | 93.1 (70.4 to 98.4) |
|  | 7-13 | 6 | 3,810 | 93.6 (85.0 to 97.2) | 2 | 3,810 | 96.0 (83.0 to 99.0) |
|  | 14-34 | 31 | 25,524 | 95.1 (92.5 to 96.8) | 10 | 25,524 | 97.0 (93.9 to 98.5) |
|  | 35-69 | 263 | 67,132 | 92.7 (90.5 to 94.4) | 101 | 67,132 | 94.6 (92.0 to 96.3) |
|  | 70-104 | 423 | 34,337 | 90.6 (87.9 to 92.7) | 166 | 34,337 | 92.9 (89.7 to 95.1) |
|  | 105+ | 185 | 7,009 | 86.2 (81.2 to 89.8) | 79 | 7,009 | 88.6 (82.1 to 92.7) |
|  | **Delta** | | | | | | |
|  | **Interval (days)** | **cases** | **controls** | **VE (95% CI)** | **cases** | **controls** | **VE (95% CI)** |
| **unvaccinated** | | **701** | **4,552** |  | **458** | **4,552** |  |
| Dose 1 | 0-27 | 15 | 487 | 67.2 (43.4 to 80.9) | 8 | 487 | 70.8 (39.7 to 85.8) |
|  | 28+ | 78 | 4,460 | 83.3 (78.5 to 87.0) | 40 | 4,460 | 86.8 (81.4 to 90.6) |
| Dose 2 | 0-13 | 7 | 3164 | 88.9 (76.1 to 94.9) | 5 | 3164 | 86.7 (66.9 to 94.7) |
|  | 14-174 | 2,742 | 143,293 | 90.8 (89.8 to 91.7) | 1,634 | 143,293 | 91.5 (90.4 to 92.5) |
|  | 175+ | 1,417 | 38,227 | 87.6 (86.1 to 88.9) | 826 | 38,227 | 89.1 (87.4 to 90.5) |
| Booster | 0-6 | 168 | 9,216 | 93.2 (91.7 to 94.3) | 94 | 9,216 | 94.3 (92.7 to 95.5) |
|  | 7-13 | 57 | 10,883 | 97.9 (97.3 to 98.5) | 29 | 10,883 | 98.4 (97.6 to 98.9) |
|  | 14-34 | 63 | 38,663 | 99.2 (98.9 to 99.4) | 38 | 38,663 | 99.2 (98.9 to 99.5) |
|  | 35-69 | 68 | 63,758 | 99.0 (98.6 to 99.2) | 31 | 63,758 | 99.3 (98.9 to 99.5) |
|  | 70-104 | 15 | 17,461 | 98.2 (96.8 to 99.0) | 7 | 17,461 | 98.7 (97.1 to 99.4) |
|  | 105+ | 0 | 65 |  | 0 | 65 |  |

### Table S9: Vaccine effectiveness using secondary care hospital admission data (SUS) by the Delta and Omicron variants in individuals 18-64 years of age

|  |  | **Omicron** | | | **Delta** | | |
| --- | --- | --- | --- | --- | --- | --- | --- |
|  |  | **SUS Pillar1 and 2 (0 days stay in hospital)** | | | | | |
| **Interval (days)** | | **cases** | **controls** | **VE (95% CI)** | **cases** | **controls** | **VE (95% CI)** |
| **unvaccinated** | | **914** | **321** |  | **3,021** | **1,444** |  |
| Dose 1 | 0-27 | 24 | 11 | 21.9 (-89.7 to 67.9) | 197 | 149 | 35.8 (14.8 to 51.6) |
|  | 28+ | 216 | 94 | 25.0 (-9.1 to 48.5) | 486 | 756 | 65.1 (59.0 to 70.3) |
| Dose 2 | 0-13 | 18 | 11 | 63.6 (-0.4 to 86.8) | 71 | 222 | 83.4 (76.9 to 88.1) |
|  | 14-174 | 443 | 318 | 46.9 (30.5 to 59.5) | 1,604 | 3,519 | 82.5 (80.3 to 84.4) |
|  | 175+ | 645 | 434 | 41.7 (25.3 to 54.5) | 326 | 843 | 75.0 (69.7 to 79.3) |
| Booster | 0-6 | 75 | 83 | 62.6 (40.1 to 76.7) | 25 | 135 | 86.7 (78.2 to 91.8) |
|  | 7-13 | 81 | 90 | 75.3 (61.1 to 84.3) | 8 | 125 | 92.5 (83.9 to 96.5) |
|  | 14-34 | 368 | 301 | 72.7 (63.9 to 79.3) | 13 | 306 | 95.3 (91.4 to 97.4) |
|  | 35-69 | 949 | 434 | 62.6 (52.0 to 70.9) | 10 | 240 | 94.0 (88.0 to 97.0) |
|  | 70-104 | 1,197 | 194 | 44.8 (26.1 to 58.8) | 5 | 66 | 71.8 (21.2 to 89.9) |
|  | 105+ | 696 | 42 | 11.7 (-36.5 to 42.9) | 0 | 3 |  |
|  |  | **SUS Pillar1 and 2 ARI code not in primary diagnostic field but 1+ days stay** | | | | | |
|  |  | **cases** | **controls** | **VE (95% CI)** | **cases** | **controls** | **VE (95% CI)** |
| **unvaccinated** | | **2,472** | **1,133** |  | **5,032** | **5,625** |  |
| Dose 1 | 0-27 | 64 | 32 | 0.0 (-92.6 to 48.0) | 200 | 424 | 40.3 (24.8 to 52.6) |
|  | 28+ | 530 | 339 | 16.2 (-3.7 to 32.3) | 876 | 3,634 | 54.7 (49.5 to 59.3) |
| Dose 2 | 0-13 | 26 | 14 | 8.3 (-152.9 to 66.7) | 102 | 778 | 63.2 (51.6 to 72.0) |
|  | 14-174 | 841 | 666 | 29.5 (15.1 to 41.5) | 2602 | 13,340 | 71.1 (68.7 to 73.2) |
|  | 175+ | 1,252 | 1,536 | 17.8 (4.4 to 29.3) | 486 | 3,032 | 61.8 (56.1 to 66.7) |
| Booster | 0-6 | 83 | 180 | 19.8 (-25.2 to 48.6) | 34 | 343 | 76.7 (64.6 to 84.7) |
|  | 7-13 | 88 | 279 | 58.5 (39.3 to 71.6) | 17 | 431 | 86.4 (76.9 to 92.1) |
|  | 14-34 | 512 | 884 | 56.2 (47.3 to 63.7) | 30 | 980 | 87.3 (80.9 to 91.6) |
|  | 35-69 | 960 | 1,464 | 56.6 (49.6 to 62.7) | 16 | 844 | 87.4 (77.4 to 93.0) |
|  | 70-104 | 490 | 619 | 50.2 (40.0 to 58.7) | 0 | 168 |  |
|  | 105+ | 151 | 162 | 50.9 (34.3 to 63.3) | 0 | 2 |  |
|  |  | **SUS Pillar1 and 2 at least 1 day stay and ARI code in primary diagnostic field** | | | | | |
|  |  | **cases** | **controls** | **VE (95% CI)** | **cases** | **controls** | **VE (95% CI)** |
| **unvaccinated** | | **1,307** | **865** |  | **20,009** | **4,235** |  |
| Dose 1 | 0-27 | 29 | 33 | 40.3 (-18 to 69.8) | 755 | 356 | 44.7 (34.3 to 53.5) |
|  | 28+ | 237 | 337 | 42.8 (26.3 to 55.5) | 1560 | 2,896 | 82.7 (81.0 to 84.3) |
| Dose 2 | 0-13 | 7 | 19 | 87.5 (59.5 to 96.1) | 192 | 725 | 90.5 (88.1 to 92.5) |
|  | 14-174 | 214 | 596 | 71.6 (63.4 to 77.9) | 7571 | 12,237 | 91.0 (90.4 to 91.5) |
|  | 175+ | 705 | 1,385 | 52.5 (43.3 to 60.1) | 1946 | 2,905 | 84.2 (82.5 to 85.7) |
| Booster | 0-6 | 45 | 189 | 70.1 (51.8 to 81.4) | 164 | 360 | 88.9 (85.7 to 91.5) |
|  | 7-13 | 40 | 313 | 87.7 (79.9 to 92.5) | 65 | 484 | 95.0 (93.1 to 96.4) |
|  | 14-34 | 194 | 921 | 87.8 (84.3 to 90.5) | 66 | 1,079 | 97.5 (96.7 to 98.2) |
|  | 35-69 | 504 | 1,389 | 83.4 (80 to 86.2) | 53 | 903 | 96.0 (94.3 to 97.2) |
|  | 70-104 | 509 | 690 | 76.3 (70.8 to 80.7) | 8 | 191 | 89.7 (75.9 to 95.6) |
|  | 105+ | 189 | 149 | 66.3 (53.6 to 75.5) | 0 | 1 |  |
|  |  | **SUS Pillar1 and 2 at least 2 days stay and ARI code in primary diagnostic field** | | | | | |
|  |  | **cases** | **controls** | **VE (95% CI)** | **cases** | **controls** | **VE (95% CI)** |
| **unvaccinated** | | **1,016** | **696** |  | **17836,** | **3,473** |  |
| Dose 1 | 0-27 | 25 | 24 | 36.2 (-33.9 to 69.6) | 633 | 270 | 43.5 (31.1 to 53.6) |
|  | 28+ | 178 | 272 | 44.1 (25.6 to 58.0) | 1,326 | 2,410 | 82.7 (80.8 to 84.4) |
| Dose 2 | 0-13 | 5 | 15 | 88.9 (58.4 to 97.0) | 157 | 605 | 90.8 (88.0 to 92.9) |
|  | 14-174 | 135 | 430 | 69.0 (58.1 to 77.0) | 6,543 | 10,137 | 91.1 (90.5 to 91.7) |
|  | 175+ | 517 | 1,150 | 56.1 (46.4 to 64.0) | 1,732 | 2,438 | 84.5 (82.7 to 86.1) |
| Booster | 0-6 | 30 | 147 | 74.3 (55.9 to 85.0) | 137 | 292 | 90.0 (86.7 to 92.5) |
|  | 7-13 | 23 | 252 | 90.9 (83.2 to 95.1) | 58 | 394 | 94.8 (92.6 to 96.3) |
|  | 14-34 | 134 | 762 | 88.6 (84.9 to 91.5) | 63 | 908 | 97.4 (96.4 to 98.1) |
|  | 35-69 | 330 | 1,133 | 85.8 (82.4 to 88.5) | 47 | 745 | 96.1 (94.3 to 97.3) |
|  | 70-104 | 310 | 567 | 80.2 (74.9 to 84.4) | 5 | 161 | 91.3 (75.4 to 96.9) |
|  | 105+ | 124 | 120 | 67.4 (53.1 to 77.4) | 0 | 1 |  |
|  |  | **SUS Pillar1 and 2 at least 3 days stay and ARI code in primary diagnostic field** | | | | | |
|  |  | **cases** | **controls** | **VE (95% CI)** | **cases** | **controls** | **VE (95% CI)** |
| **unvaccinated** | | **804** | **560** |  | **15819** | **2,863** |  |
| Dose 1 | 0-27 | 20 | 19 | 40.0 (-40.4 to 74.4) | 525 | 213 | 46.3 (32.8 to 57.1) |
|  | 28+ | 133 | 234 | 51.5 (33.2 to 64.8) | 1129 | 2,039 | 83.0 (80.9 to 84.8) |
| Dose 2 | 0-13 | 5 | 12 | 87.6 (53.7 to 96.7) | 134 | 500 | 91.1 (88.2 to 93.3) |
|  | 14-174 | 91 | 338 | 72.7 (61.4 to 80.7) | 5738 | 8,405 | 91.0 (90.3 to 91.7) |
|  | 175+ | 391 | 984 | 60.6 (50.7 to 68.5) | 1541 | 2,074 | 85.2 (83.3 to 86.8) |
| Booster | 0-6 | 26 | 122 | 77.2 (59.5 to 87.2) | 123 | 240 | 90.3 (86.8 to 92.9) |
|  | 7-13 | 13 | 212 | 95.0 (89.1 to 97.7) | 52 | 334 | 94.7 (92.4 to 96.4) |
|  | 14-34 | 96 | 637 | 89.8 (85.9 to 92.6) | 55 | 766 | 97.6 (96.6 to 98.3) |
|  | 35-69 | 230 | 937 | 87.8 (84.3 to 90.4) | 42 | 621 | 96.0 (94.1 to 97.4) |
|  | 70-104 | 222 | 463 | 80.4 (74.5 to 85.0) | 5 | 137 | 91.7 (76.0 to 97.1) |
|  | 105+ | 79 | 88 | 68.6 (52.3 to 79.4) | 0 | 1 |  |
|  |  | **SUS Pillar1 and 2 at least 2 days stay & oxygen with ARI code in primary diagnostic field** | | | | | |
|  |  | **cases** | **controls** | **VE (95% CI)** | **cases** | **controls** | **VE (95% CI)** |
| **unvaccinated** | | **126** | **42** |  | **3,918** | **258** |  |
| Dose 1 | 0-27 | 1 | 1 |  | 119 | 19 | 41.4 (-14.8 to 70.1) |
|  | 28+ | 9 | 18 | 87.5 (55.6 to 96.5) | 221 | 154 | 83.3 (76.7 to 88.0) |
| Dose 2 | 0-13 | 0 | 1 |  | 39 | 30 | 80.0 (55.7 to 91.0) |
|  | 14-174 | 6 | 18 | 79.1 (-36.9 to 96.8) | 1184 | 599 | 91.4 (89.2 to 93.1) |
|  | 175+ | 30 | 81 | 80.5 (48.7 to 92.6) | 322 | 175 | 86.1 (80.3 to 90.2) |
| Booster | 0-6 | 4 | 7 |  | 21 | 17 | 94.0 (83.9 to 97.8) |
|  | 7-13 | 0 | 13 |  | 8 | 18 | 95.0 (81.4 to 98.6) |
|  | 14-34 | 6 | 43 | 94.2 (76.6 to 98.6) | 13 | 50 | 96.0 (90.4 to 98.3) |
|  | 35-69 | 15 | 75 | 93.9 (81.6 to 97.9) | 18 | 55 | 93.2 (83.4 to 97.3) |
|  | 70-104 | 19 | 30 | 94 (78.8 to 98.3) | 3 | 10 |  |
|  | 105+ | 6 | 7 | 80.4 (-36.3 to 97.2) |  |  |  |
|  |  | **SUS Pillar1 and 2 at least 2 days stay & either oxygen, ventilation or ICU with ARI code in primary diagnostic field** | | | | | |
|  |  | **cases** | **controls** | **VE (95% CI)** | **cases** | **controls** | **VE (95% CI)** |
| **unvaccinated** | | **227** | **87** |  | **6,511** | **505** |  |
| Dose 1 | 0-27 | 5 | 3 |  | 174 | 34 | 51.0 (18.7 to 70.4) |
|  | 28+ | 22 | 32 | 75.0 (42.4 to 89.1) | 376 | 331 | 82.7 (78.0 to 86.4) |
| Dose 2 | 0-13 | 0 | 3 |  | 52 | 73 | 85.0 (73.0 to 91.7) |
|  | 14-174 | 12 | 47 | 86.7 (63.6 to 95.1) | 2,090 | 1,338 | 90.5 (88.9 to 91.9) |
|  | 175+ | 67 | 172 | 82.3 (67.7 to 90.3) | 583 | 360 | 84.3 (79.9 to 87.7) |
| Booster | 0-6 | 4 | 17 | 90.7 (56 to 98.1) | 45 | 42 | 92.7 (86.1 to 96.1) |
|  | 7-13 | 0 | 28 |  | 18 | 51 | 93.9 (87.2 to 97.1) |
|  | 14-34 | 9 | 107 | 97.1 (92.2 to 98.9) | 23 | 118 | 95.2 (90.7 to 97.5) |
|  | 35-69 | 31 | 155 | 94.3 (88.9 to 97.1) | 22 | 107 | 93.5 (86.2 to 97.0) |
|  | 70-104 | 36 | 66 | 89.9 (78.3 to 95.3) | 4 | 21 |  |
|  | 105+ | 11 | 12 | 75.9 (15.8 to 93.1) |  |  |  |

### Table S10: Vaccine effectiveness using secondary care hospital admission data (SUS) by the Delta and Omicron variants in individuals 65 years of age and older

|  |  | **Omicron** | | | **Delta** | | |
| --- | --- | --- | --- | --- | --- | --- | --- |
|  |  | **SUS Pillar1 and 2 (0 days stay in hospital)** | | | | | |
|  | **Interval (days)** | **cases** | **controls** | **VE (95% CI)** | **cases** | **controls** | **VE (95% CI)** |
| **unvaccinated** | | **69** | **60** |  | **104** | **202** |  |
| Dose 1 | 0-27 | 1 | 6 |  | 4 | 17 |  |
|  | 28+ | 14 | 25 | 67.5 (6.7 to 88.6) | 20 | 214 | 65.4 (37.0 to 81.0) |
| Dose 2 | 0-13 |  |  |  | 2 | 95 | 66.8 (-64.5 to 93.3) |
|  | 14-174 | 6 | 13 | 55.1 (-131.8 to 91.3) | 521 | 3,240 | 76.0 (67.0 to 82.6) |
|  | 175+ | 97 | 196 | 39.2 (-6.7 to 65.4) | 266 | 949 | 56.6 (38.8 to 69.2) |
| Booster | 0-6 | 3 | 43 | 85.6 (6.4 to 97.8) | 22 | 165 | 81.8 (66.9 to 90.0) |
|  | 7-13 | 5 | 36 | 69.2 (-18.7 to 92) | 18 | 135 | 77.8 (58.6 to 88.1) |
|  | 14-34 | 25 | 192 | 87.4 (72.5 to 94.2) | 21 | 481 | 90.8 (83.7 to 94.8) |
|  | 35-69 | 211 | 741 | 79.3 (65.7 to 87.5) | 11 | 704 | 90.9 (81.1 to 95.6) |
|  | 70-104 | 551 | 565 | 67.2 (46.6 to 79.8) | 0 | 245 |  |
|  | 105+ | 411 | 244 | 59.0 (30.5 to 75.8) | 0 | 3 |  |
|  |  | **SUS Pillar1 and 2 ARI code not in primary diagnostic field but 1+ days stay** | | | | | |
|  | **Interval (days)** | **cases** | **controls** | **VE (95% CI)** | **cases** | **controls** | **VE (95% CI)** |
| **unvaccinated** | | **354** | **636** |  | **479** | **3,194** |  |
| Dose 1 | 0-27 | 12 | 17 |  | 21 | 161 |  |
|  | 28+ | 82 | 228 | 33.2 (5.2 to 53.0) | 174 | 3,450 | 40.3 (27.4 to 50.9) |
| Dose 2 | 0-13 | 0 | 15 |  | 13 | 1,339 | 48.3 (6.8 to 71.4) |
|  | 14-174 | 35 | 273 | 66.5 (47.1 to 78.7) | 2,765 | 43,778 | 57.7 (52.4 to 62.4) |
|  | 175+ | 595 | 2,399 | 23.5 (6.4 to 37.5) | 1,535 | 13,500 | 46.7 (39.3 to 53.1) |
| Booster | 0-6 | 31 | 388 | 38.9 (-2.6 to 63.6) | 141 | 1,575 | 54.4 (43.1 to 63.5) |
|  | 7-13 | 33 | 496 | 62.5 (40.7 to 76.4) | 105 | 1,686 | 71.3 (63.2 to 77.7) |
|  | 14-34 | 179 | 2,267 | 69.3 (60.8 to 76.0) | 140 | 5,188 | 83.9 (80.0 to 87.1) |
|  | 35-69 | 805 | 6,378 | 67.2 (60.5 to 72.8) | 68 | 6,759 | 85.4 (80.5 to 89.1) |
|  | 70-104 | 1,718 | 5,148 | 59.4 (51.5 to 66.0) | 5 | 2,311 | 55.7 (-23.9 to 84.2) |
|  | 105+ | 860 | 1,822 | 56.3 (46.9 to 64.0) | 0 | 22 |  |
|  |  | **SUS Pillar1 and 2 at least 1 day stay and ARI code in primary diagnostic field** | | | | | |
|  | **Interval (days)** | **cases** | **controls** | **VE (95% CI)** | **cases** | **controls** | **VE (95% CI)** |
| **unvaccinated** | | **966** | **736** |  | **4,374** | **3,197** |  |
| Dose 1 | 0-27 | 18 | 30 | 57.4 (-0.7 to 82) | 79 | 200 | 55.1 (35.7 to 68.7) |
|  | 28+ | 146 | 274 | 52.3 (35.8 to 64.5) | 623 | 3,522 | 74.7 (71.6 to 77.6) |
| Dose 2 | 0-13 | 1 | 7 |  | 41 | 1,373 | 82.8 (75.0 to 88.2) |
|  | 14-174 | 74 | 326 | 80.5 (72.2 to 86.3) | 10,252 | 46,368 | 85.7 (84.7 to 86.6) |
|  | 175+ | 972 | 2,986 | 58.4 (51.0 to 64.7) | 6,299 | 15,711 | 78.2 (76.6 to 79.8) |
| Booster | 0-6 | 42 | 562 | 78.5 (66.8 to 86.1) | 512 | 2,109 | 86.8 (84.8 to 88.5) |
|  | 7-13 | 44 | 637 | 82.2 (73.0 to 88.3) | 401 | 2,200 | 88.0 (86.2 to 89.6) |
|  | 14-34 | 189 | 2,949 | 91.3 (89.1 to 93.0) | 354 | 6,794 | 96.4 (95.8 to 96.9) |
|  | 35-69 | 892 | 8,390 | 88.9 (87.1 to 90.6) | 208 | 9,008 | 95.7 (94.9 to 96.4) |
|  | 70-104 | 1,826 | 6,514 | 87.6 (85.6 to 89.3) | 18 | 3,187 | 91.2 (85.2 to 94.8) |
|  | 105+ | 983 | 2,332 | 84.1 (81.2 to 86.5) | 0 | 26 |  |
|  |  | **SUS Pillar1 and 2 at least 2 days stay and ARI code in primary diagnostic field** | | | | | |
|  | **Interval (days)** | **cases** | **controls** | **VE (95% CI)** | **cases** | **controls** | **VE (95% CI)** |
| **unvaccinated** | | **894** | **667** |  | **4,198** | **2,976** |  |
| Dose 1 | 0-27 | 17 | 27 | 43.9 (-41 to 77.7) | 73 | 191 | 59.2 (41.1 to 71.8) |
|  | 28+ | 134 | 248 | 53.4 (36.3 to 65.9) | 593 | 3,263 | 74.5 (71.2 to 77.4) |
| Dose 2 | 0-13 | 0 | 5 |  | 40 | 1,253 | 81.8 (73.3 to 87.6) |
|  | 14-174 | 65 | 305 | 82.3 (74.3 to 87.8) | 9,584 | 42,489 | 85.6 (84.6 to 86.6) |
|  | 175+ | 913 | 2,733 | 57.7 (49.6 to 64.4) | 5,937 | 14,482 | 78.3 (76.6 to 79.9) |
| Booster | 0-6 | 41 | 507 | 77.9 (65.3 to 85.9) | 470 | 1,893 | 86.9 (84.8 to 88.6) |
|  | 7-13 | 39 | 595 | 84.7 (76.0 to 90.2) | 362 | 2,024 | 88.6 (86.8 to 90.1) |
|  | 14-34 | 177 | 2,693 | 91.3 (89.1 to 93.1) | 339 | 6,162 | 96.3 (95.7 to 96.8) |
|  | 35-69 | 801 | 7,616 | 89.3 (87.3 to 90.9) | 194 | 8,199 | 95.7 (94.8 to 96.4) |
|  | 70-104 | 1,637 | 5,825 | 88.1 (86.1 to 89.9) | 18 | 2,849 | 90.2 (83.5 to 94.3) |
|  | 105+ | 847 | 2,036 | 85.3 (82.4 to 87.6) | 0 | 25 |  |
|  |  | **SUS Pillar1 and 2 at least 3 days stay and ARI code in primary diagnostic field** | | | | | |
|  | **Interval (days)** | **cases** | **controls** | **VE (95% CI)** | **cases** | **controls** | **VE (95% CI)** |
| **unvaccinated** | | **835** | **603** |  | **3,947** | **2,753** |  |
| Dose 1 | 0-27 | 15 | 23 | 35.1 (-80.6 to 76.7) | 70 | 166 | 55.0 (33.8 to 69.4) |
|  | 28+ | 120 | 232 | 57.6 (41.3 to 69.3) | 559 | 3,000 | 74.2 (70.7 to 77.3) |
| Dose 2 | 0-13 | 0 | 4 |  | 36 | 1,152 | 82.7 (74.0 to 88.5) |
|  | 14-174 | 61 | 281 | 82.2 (73.7 to 87.9) | 8,815 | 38,698 | 85.7 (84.7 to 86.7) |
|  | 175+ | 841 | 2,488 | 57.8 (49.4 to 64.9) | 5,561 | 13,261 | 78.1 (76.3 to 79.7) |
| Booster | 0-6 | 38 | 463 | 77.9 (64.7 to 86.2) | 436 | 1,702 | 86.4 (84.2 to 88.3) |
|  | 7-13 | 38 | 552 | 84.5 (75.5 to 90.2) | 335 | 1,831 | 88.3 (86.4 to 90.0) |
|  | 14-34 | 163 | 2,416 | 91.4 (89.0 to 93.2) | 317 | 5,536 | 96.1 (95.5 to 96.6) |
|  | 35-69 | 715 | 6,772 | 89.5 (87.5 to 91.2) | 179 | 7,331 | 95.6 (94.7 to 96.3) |
|  | 70-104 | 1,469 | 5,191 | 88.6 (86.5 to 90.3) | 16 | 2,546 | 89.7 (82.1 to 94.1) |
|  | 105+ | 724 | 1,771 | 86.4 (83.6 to 88.7) | 0 | 22 |  |
|  |  | **SUS Pillar1 and 2 at least 2 days stay & oxygen with ARI code in primary diagnostic field** | | | | | |
|  | **Interval (days)** | **cases** | **controls** | **VE (95% CI)** | **cases** | **controls** | **VE (95% CI)** |
| **unvaccinated** | | **138** | **41** |  | **920** | **163** |  |
| Dose 1 | 0-27 | 2 | 1 |  | 18 | 6 |  |
|  | 28+ | 10 | 6 | 48.3 (-169.1 to 90) | 71 | 124 | 82.3 (71.7 to 89.0) |
| Dose 2 | 0-13 |  |  |  | 6 | 62 | 88.1 (64.4 to 96.0) |
|  | 14-174 | 5 | 13 | 87.7 (50.9 to 96.9) | 1,503 | 1,940 | 90.1 (87.3 to 92.2) |
|  | 175+ | 69 | 154 | 74.0 (47.6 to 87.1) | 893 | 694 | 83.9 (78.8 to 87.8) |
| Booster | 0-6 | 6 | 27 | 70.0 (-33.9 to 93.3) | 69 | 106 | 93.5 (89.2 to 96.0) |
|  | 7-13 | 3 | 31 | 86.7 (-13.9 to 98.5) | 48 | 102 | 91.6 (86.1 to 94.9) |
|  | 14-34 | 10 | 125 | 95.9 (89.0 to 98.4) | 56 | 318 | 97.7 (96.3 to 98.5) |
|  | 35-69 | 70 | 398 | 93.9 (88.4 to 96.8) | 39 | 446 | 96.7 (94.4 to 98.1) |
|  | 70-104 | 146 | 281 | 93.2 (87.5 to 96.2) | 2 | 131 | 90.0 (43.4 to 98.2) |
|  | 105+ | 58 | 78 | 90.1 (79.7 to 95.2) |  |  |  |
|  |  | **SUS Pillar1 and 2 at least 2 days stay & either oxygen, ventilation or ICU with ARI code in primary diagnostic field** | | | | | |
|  | **Interval (days)** | **cases** | **controls** | **VE (95% CI)** | **cases** | **controls** | **VE (95% CI)** |
| **unvaccinated** | | **215** | **70** |  | **1664** | **294** |  |
| Dose 1 | 0-27 | 4 | 3 |  | 27 | 14 | 24.5 (-141.3 to 76.4) |
|  | 28+ | 16 | 16 | 78.3 (43.7 to 91.7) | 131 | 269 | 86.7 (81.4 to 90.5) |
| Dose 2 | 0-13 |  |  |  | 10 | 112 | 89.8 (74.5 to 95.9) |
|  | 14-174 | 7 | 30 | 90.9 (72.6 to 97.0) | 2570 | 3,491 | 90.5 (88.6 to 92.1) |
|  | 175+ | 108 | 249 | 73.4 (55.1 to 84.3) | 1494 | 1,174 | 84.9 (81.5 to 87.7) |
| Booster | 0-6 | 6 | 37 | 89.2 (63.1 to 96.8) | 122 | 150 | 92.1 (88.4 to 94.6) |
|  | 7-13 | 4 | 57 | 94.7 (71.6 to 99.0) | 92 | 184 | 92.3 (88.9 to 94.7) |
|  | 14-34 | 17 | 236 | 95.8 (91.3 to 97.9) | 89 | 543 | 97.5 (96.5 to 98.3) |
|  | 35-69 | 115 | 638 | 92.8 (88.4 to 95.6) | 61 | 692 | 96.1 (94.0 to 97.4) |
|  | 70-104 | 220 | 467 | 92.5 (88.1 to 95.2) | 5 | 223 | 87.0 (58.6 to 95.9) |
|  | 105+ | 95 | 118 | 86.8 (77.1 to 92.3) |  |  |  |

### Table S11: Vaccine effectiveness by manufacturer against hospital admissions from emergency care (ECDS) within 14 days of the test date by the Omicron variant in symptomatic individuals 18 to 64 years of age

| **Manufacturer** | **Doses** |  | **Pillar 2 symptomatic with ECDS admission within 14 days** | | | **Pillar 2 symptomatic with ARI coded ECDS admission within 14 days** | | |
| --- | --- | --- | --- | --- | --- | --- | --- | --- |
|  |  | **Interval (days)** | **cases** | **controls** | **VE (95% CI)** | **cases** | **controls** | **VE** |
|  |  | **unvaccinated** | **643** | **127,803** |  | **171** | **127803** |  |
| ChAdOx1-S | Dose 1 | 0-27 | 0 | 31 |  | 0 | 31 |  |
|  |  | 28+ | 50 | 9267 | 17.6 (-10.5 to 38.5) | 14 | 9267 | 30.3 (-21.4 to 60.0) |
|  | Dose 2 | 0-13 | 0 | 223 |  | 0 | 223 |  |
|  |  | 14-174 | 66 | 76181 | 38.5 (20.2 to 52.6) | 15 | 76181 | 52.6 (18.7 to 72.4) |
|  |  | 175+ | 444 | 151244 | 31.6 (22.3 to 39.7) | 102 | 151244 | 50.0 (35.4 to 61.3) |
|  | Booster (any) | 0-6 | 62 | 51422 | 58.2 (45.4 to 68.0) | 12 | 51422 | 74.1 (52.9 to 85.8) |
|  | Booster (BNT162b2) | 7-13 | 25 | 37798 | 85.2 (77.9 to 90.1) | 9 | 37798 | 83.0 (66.4 to 91.4) |
|  |  | 14-34 | 121 | 110195 | 84.1 (80.5 to 87.0) | 25 | 110195 | 89.7 (84.1 to 93.3) |
|  |  | 35-69 | 278 | 114444 | 76.3 (72.3 to 79.6) | 53 | 114444 | 87.5 (82.5 to 91.0) |
|  |  | 70+ | 161 | 32156 | 65.6 (58.5 to 71.6) | 51 | 32156 | 73.4 (62.3 to 81.2) |
|  |  | 105+ | 52 | 4808 | 38.2 (16.0 to 54.5) | 18 | 4808 |  |
|  | Booster (mRNA-1273) | 7-13 | 11 | 21600 | 88.1 (78.3 to 93.5) | 0 | 21600 |  |
|  |  | 14-34 | 50 | 56829 | 88.3 (84.3 to 91.3) | 5 | 56829 | 96.2 (90.7 to 98.4) |
|  |  | 35-69 | 83 | 43238 | 80.7 (75.5 to 84.8) | 16 | 43238 | 88.1 (79.8 to 93.0) |
|  |  | 70+ | 15 | 5316 | 74.1 (56.3 to 84.7) | 1 | 5316 |  |
| BNT162b2 | Dose 1 | 0-27 | 25 | 8273 | 38.0 (7.3 to 58.5) | 6 | 8273 | 41.7 (-32.0 to 74.3) |
|  |  | 28+ | 88 | 35531 | 44.9 (31.0 to 56.0) | 12 | 35531 | 70.4 (46.8 to 83.6) |
|  | Dose 2 | 0-13 | 13 | 6607 | 52.8 (18.1 to 72.8) | 3 | 6607 | 56.8 (-35.8 to 86.2) |
|  |  | 14-174 | 340 | 321184 | 61.0 (55.3 to 65.9) | 59 | 321184 | 71.7 (61.6 to 79.2) |
|  |  | 175+ | 207 | 60035 | 33.0 (21.3 to 42.9) | 48 | 60035 | 48.0 (27.8 to 62.5) |
|  | Booster (any) | 0-6 | 41 | 43987 | 73.7 (63.8 to 80.9) | 5 | 43987 | 87.2 (68.7 to 94.8) |
|  | Booster (BNT162b2) | 7-13 | 37 | 29861 | 77.5 (68.4 to 83.9) | 4 | 29861 | 92.5 (76.4 to 97.6) |
|  |  | 14-34 | 84 | 77362 | 82.8 (78.4 to 86.3) | 17 | 77362 | 87.4 (79.2 to 92.4) |
|  |  | 35-69 | 183 | 119375 | 78.6 (74.6 to 82.0) | 41 | 119375 | 86.1 (80.1 to 90.2) |
|  |  | 70+ | 188 | 67343 | 72.5 (67.1 to 77.0) | 46 | 67343 | 81.8 (74.0 to 87.3) |
|  |  | 105+ | 79 | 21177 | 67.7 (58.2 to 75.0) | 19 | 21177 |  |
|  | Booster (mRNA-1273) | 7-13 | 8 | 12923 | 88.5 (76.8 to 94.3) | 2 | 12923 | 88.4 (53.2 to 97.1) |
|  |  | 14-34 | 28 | 29200 | 87.3 (81.2 to 91.3) | 5 | 29200 | 91.0 (78.1 to 96.3) |
|  |  | 35-69 | 32 | 23649 | 84.5 (77.9 to 89.2) | 4 | 23649 | 93.3 (81.9 to 97.5) |
|  |  | 70+ | 11 | 2144 | 60.6 (27.7 to 78.5) | 1 | 2144 | 0 |

### Table S12: Vaccine effectiveness by manufacturer against hospital admissions from emergency care (ECDS) within 14 days of the test date by the Omicron variant in symptomatic individuals 65 years of age and older

| **Manufacturer** |  |  | **Pillar 2 symptomatic with ECDS admission within 14 days** | | | **Pillar 2 symptomatic with ARI coded ECDS admission within 14 days** | | |
| --- | --- | --- | --- | --- | --- | --- | --- | --- |
|  | **Doses** | **Interval (days)** | **cases** | **controls** | **VE (95% CI)** | **cases** | **controls** | **VE (95% CI)** |
|  |  | **unvaccinated** | **103** | **1,705** |  | **49** | **1,705** |  |
| **ChAdOx1-S** | Dose 1 | 0-27 | 0 | 2 |  | 0 | 2 |  |
|  |  | 28+ | 10 | 280 |  | 4 | 280 |  |
|  | Dose 2 | 0-13 | 0 | 6 |  | 0 | 6 |  |
|  |  | 14-174 | 5 | 390 | 55.3 (-20.1 to 83.3) | 2 | 390 | 69.9 (-38.4 to 93.5) |
|  |  | 175+ | 88 | 4,456 | 56.3 (39.6 to 68.3) | 35 | 4,456 | 68.9 (49.7 to 80.8) |
|  | Booster (any) | 0-6 | 5 | 1,554 | 84.8 (61.1 to 94.1) | 1 | 1,554 | 95.1 (62.8 to 99.4) |
|  | Booster (BNT162b2) | 7-13 | 3 | 1,872 | 93.0 (77.5 to 97.8) | 1 | 1,872 | 95.5 (67.0 to 99.4) |
|  |  | 14-34 | 21 | 14,166 | 93.5 (89.3 to 96.0) | 8 | 14,166 | 95.3 (89.7 to 97.8) |
|  |  | 35-69 | 137 | 35,872 | 92.7 (90.3 to 94.6) | 47 | 35,872 | 95.1 (92.3 to 96.8) |
|  |  | 70+ | 154 | 14,449 | 91.0 (88.1 to 93.3) | 60 | 14,449 | 93.3 (89.7 to 95.6) |
|  |  | 105+ | 71 | 2,602 | 82.9 (75.4 to 88.1) | 30 | 2,602 | 85.1 (74.4 to 91.4) |
|  | Booster (mRNA-1273) | 7-13 | 0 | 811 |  | 0 | 811 |  |
|  |  | 14-34 | 2 | 3,472 | 98.1 (92.1 to 99.5) | 1 | 3,472 | 98.1 (85.8 to 99.7) |
|  |  | 35-69 | 10 | 3,445 | 94.9 (90.0 to 97.4) | 1 | 3,445 | 98.9 (92.1 to 99.9) |
|  |  | 70+ | 5 | 775 | 89.6 (73.8 to 95.9) | 3 | 775 |  |
| **BNT162b2** | Dose 1 | 0-27 | 2 | 151 | 76.1 (-8.9 to 94.7) | 1 | 151 |  |
|  |  | 28+ | 6 | 324 | 78.0 (47.7 to 90.8) | 1 | 324 | 93 (47.8 to 99.1) |
|  | Dose 2 | 0-13 | 0 | 22 |  | 0 | 22 |  |
|  |  | 14-174 | 8 | 569 | 85.8 (69.4 to 93.4) | 3 | 569 | 90.4 (67.1 to 97.2) |
|  |  | 175+ | 28 | 1,792 | 75.6 (61.4 to 84.6) | 7 | 1,792 | 89.5 (75.8 to 95.4) |
|  | Booster (any) | 0-6 | 4 | 569 | 72.7 (19.1 to 90.8) | 1 | 569 |  |
|  | Booster (BNT162b2) | 7-13 | 2 | 882 | 90.1 (58.6 to 97.6) | 1 | 882 |  |
|  |  | 14-34 | 4 | 6,805 | 97.4 (92.8 to 99.1) | 0 | 6,805 |  |
|  |  | 35-69 | 113 | 26,699 | 91.7 (88.8 to 93.9) | 51 | 26,699 | 92.8 (88.8 to 95.3) |
|  |  | 70+ | 260 | 18,863 | 90.2 (87.2 to 92.5) | 102 | 18,863 | 92.7 (89.1 to 95) |
|  |  | 105+ | 113 | 4,379 | 88.0 (83.4 to 91.4) | 48 | 4,379 | 90.3 (84.3 to 94.1) |
|  | Booster (mRNA-1273) | 7-13 | 1 | 241 | 89.0 (17.2 to 98.5) | 0 | 241 |  |
|  |  | 14-34 | 4 | 1,076 | 91.2 (75.5 to 96.9) | 1 | 1,076 | 95.9 (69.5 to 99.5) |
|  |  | 35-69 | 1 | 1,112 |  | 0 | 1,112 |  |
|  |  | 70+ | 0 | 276 |  | 0 | 276 |  |

### Table S13: Vaccine effectiveness using secondary care hospital admission data (SUS) by manufacturer for the Omicron variant in individuals 18-64 years of age

| **Manufacturer** |  |  | **SUS Pillar1 and 2 at least 2 days stay and ARI code in primary diagnostic field** | | | **SUS Pillar1 and 2 at least 2 days stay & either O2, Vent or ICU and ARI code in primary diagnostic field** | | |
| --- | --- | --- | --- | --- | --- | --- | --- | --- |
|  |  | **Interval (days)** | **cases** | **controls** | **VE (95% CI)** | **cases** | **controls** | **VE** |
|  | **Doses** | **unvaccinated** | **1016** | **696** |  | **227** | **87** |  |
| **ChAdOx1-S** | Dose 1 |  |  |  |  |  |  |  |
|  |  | 28+ | 88 | 153 | 48.5 (25.7 to 64.3) | 12 | 23 | 82.8 (54.3 to 93.5) |
|  | Dose 2 |  |  |  |  | 0 | 1 |  |
|  |  | 14-174 | 37 | 182 | 59.0 (31.9 to 75.3) | 7 | 25 |  |
|  |  | 175+ | 406 | 877 | 53.0 (41.7 to 62.0) | 55 | 140 | 81.9 (65.7 to 90.4) |
|  | Booster (any) | 0-6 | 21 | 120 | 78.0 (59.5 to 88.1) | 4 | 14 | 90.4 (53.9 to 98.0) |
|  | Booster (BNT162b2) | 7-13 | 13 | 131 | 90.2 (78.1 to 95.6) | 0 | 16 |  |
|  |  | 14-34 | 63 | 415 | 88.9 (83.8 to 92.4) | 5 | 58 | 96.6 (88.7 to 99.0) |
|  |  | 35-69 | 184 | 632 | 83.9 (79.1 to 87.5) | 19 | 88 | 93.2 (85.4 to 96.9) |
|  |  | 70+ | 151 | 327 | 82.2 (76.3 to 86.7) | 20 | 38 | 90.6 (75.7 to 96.4) |
|  |  | 105+ | 61 | 67 | 69.0 (50.3 to 80.7) | 7 | 6 |  |
|  | Booster (mRNA-1273) | 7-13 | 3 | 59 | 97.2 (86.1 to 99.4) | 0 | 3 |  |
|  |  | 14-34 | 16 | 127 | 93.0 (86.4 to 96.4) | 2 | 17 | 96.0 (64.9 to 99.5) |
|  |  | 35-69 | 32 | 123 | 89.2 (82.5 to 93.3) | 2 | 10 | 95.1 (72.9 to 99.1) |
|  |  | 70+ | 13 | 30 |  | 0 | 3 |  |
| **BNT162b2** | Dose 1 | 0-27 | 24 | 21 |  | 4 | 3 |  |
|  |  | 28+ | 89 | 110 | 31.7 (-3.1 to 54.8) | 10 | 8 |  |
|  | Dose 2 | 0-13 | 5 | 12 |  | 0 | 2 |  |
|  |  | 14-174 | 89 | 225 | 73.8 (62.5 to 81.7) | 5 | 18 | 88.7 (56.0 to 97.1) |
|  |  | 175+ | 108 | 272 | 65.1 (51.3 to 74.9) | 10 | 32 | 82.3 (45.6 to 94.2) |
|  | Booster (any) | 0-6 | 9 | 27 |  | 0 | 3 |  |
|  | Booster (BNT162b2) | 7-13 | 4 | 50 | 85.2 (47.1 to 95.8) | 0 | 8 |  |
|  |  | 14-34 | 44 | 168 | 79.7 (66.3 to 87.7) | 1 | 24 | 99.5 (90.2 to 100.0) |
|  |  | 35-69 | 100 | 335 | 86.6 (81.3 to 90.4) | 10 | 51 | 95.1 (86.7 to 98.2) |
|  |  | 70+ | 139 | 199 | 79.3 (71.3 to 85.0) | 15 | 24 | 88.0 (67.1 to 95.6) |
|  |  | 105+ | 63 | 53 | 66.0 (44.5 to 79.2) | 4 | 6 |  |
|  | Booster (mRNA-1273) | 7-13 | 2 | 9 |  | 0 | 1 |  |
|  |  | 14-34 | 10 | 50 | 94.3 (85.0 to 97.8) | 1 | 8 |  |
|  |  | 35-69 | 13 | 38 | 89.8 (77.9 to 95.3) | 0 | 5 |  |
|  |  | 70+ | 7 | 11 |  | 1 | 1 |  |

### Table S14: Vaccine effectiveness using secondary care hospital admission data (SUS) by manufacturer for the Omicron variant in individuals 65 years of age and older

| **Manufacturer** |  |  | **SUS Pillar1 and 2 at least 2 days stay and ARI code in primary diagnostic field** | | | **SUS Pillar1 and 2 at least 2 days stay & either oxygen, ventilation or ICU with ARI code in primary diagnostic field** | | |
| --- | --- | --- | --- | --- | --- | --- | --- | --- |
|  |  | **Interval (days)** | **cases** | **controls** | **VE (95% CI)** | **cases** | **controls** | **VE (95% CI)** |
|  | **Doses** | **unvaccinated** | **894** | **667** |  | **215** | **70** |  |
| **ChAdOx1-S** | Dose 1 | 0-27 | 0 | 2 |  | 0 | 1 |  |
|  |  | 28+ | 80 | 164 | 48.1 (23.5 to 64.8) | 11 | 11 | 77.3 (29.2 to 92.7) |
|  | Dose 2 | 0-13 | 0 | 1 |  |  |  |  |
|  |  | 14-174 | 35 | 151 | 71.2 (50.0 to 83.4) | 2 | 15 |  |
|  |  | 175+ | 640 | 1,815 | 53.1 (43.4 to 61.2) | 70 | 171 | 71.4 (48.6 to 84.0) |
|  | Booster (any) | 0-6 | 30 | 378 | 76.7 (61.0 to 86.1) | 4 | 28 | 92.1 (65.9 to 98.2) |
|  | Booster (BNT162b2) | 7-13 | 19 | 322 | 85.4 (73.4 to 92.0) | 2 | 29 | 94.6 (36.2 to 99.5) |
|  |  | 14-34 | 96 | 1,536 | 91.3 (88.5 to 93.5) | 12 | 144 | 95.0 (88.8 to 97.7) |
|  |  | 35-69 | 464 | 3,777 | 89.2 (87.1 to 91.0) | 68 | 315 | 93.4 (88.8 to 96.2) |
|  |  | 70+ | 752 | 2,245 | 87.6 (85.2 to 89.6) | 101 | 179 | 91.8 (86.4 to 95.1) |
|  |  | 105+ | 240 | 592 | 86.1 (82.5 to 88.9) | 25 | 44 | 92.1 (83.8 to 96.2) |
|  | Booster (mRNA-1273) | 7-13 | 5 | 66 |  | 0 | 11 |  |
|  |  | 14-34 | 22 | 223 | 92.9 (87.7 to 95.9) | 1 | 17 | 99.1 (85.8 to 99.9) |
|  |  | 35-69 | 38 | 244 | 92.7 (89.1 to 95.2) | 6 | 25 | 96.3 (88.5 to 98.8) |
|  |  | 70+ | 19 | 94 | 91.8 (85.9 to 95.3) | 1 | 7 |  |
| **BNT162b2** | Dose 1 | 0-27 | 13 | 24 | 54.8 (-25.3 to 83.7) | 3 | 2 |  |
|  |  | 28+ | 53 | 83 | 59.5 (35.5 to 74.6) | 5 | 5 |  |
|  | Dose 2 | 0-13 | 0 | 4 |  |  |  |  |
|  |  | 14-174 | 29 | 150 | 87.6 (79.4 to 92.5) | 4 | 15 | 88.9 (57.3 to 97.1) |
|  |  | 175+ | 273 | 918 | 65.4 (56.6 to 72.5) | 38 | 78 | 76.7 (52.9 to 88.5) |
|  | Booster (any) | 0-6 | 11 | 129 | 80.6 (54.4 to 91.7) | 2 | 9 |  |
|  | Booster (BNT162b2) | 7-13 | 13 | 161 | 86.4 (69.1 to 94.0) | 2 | 14 | 93.2 (10.0 to 99.5) |
|  |  | 14-34 | 51 | 822 | 90.0 (85.4 to 93.2) | 4 | 68 | 95.4 (80.9 to 98.9) |
|  |  | 35-69 | 275 | 3467 | 88.4 (85.7 to 90.6) | 39 | 288 | 90.1 (82.1 to 94.6) |
|  |  | 70+ | 863 | 3450 | 88.4 (86.2 to 90.2) | 117 | 278 | 92.8 (88.2 to 95.7) |
|  |  | 105+ | 607 | 1439 | 85.2 (82.1 to 87.7) | 70 | 74 | 83.7 (70.6 to 91.0) |
|  | Booster (mRNA-1273) | 7-13 | 2 | 45 | 92.9 (50.2 to 99.0) | 0 | 3 |  |
|  |  | 14-34 | 8 | 112 | 92.9 (83.0 to 97.1) | 0 | 7 |  |
|  |  | 35-69 | 24 | 128 | 90.9 (84.8 to 94.5) | 2 | 10 |  |
|  |  | 70+ | 3 | 41 | 97.3 (90.8 to 99.2) | 1 | 3 |  |

Table S15: SUS ICD10 and OPCS code lists

| **SUS Acute respiratory illness ICD10 code list** | |
| --- | --- |
| J04* | Acute laryngitis and tracheitis |
| J09* | Influenza due to identified avian influenza virus |
| J10* | Influenza with pneumonia, other influenza virus identified |
| J11* | Influenza with pneumonia, virus not identified |
| J12* | Viral pneumonia, not elsewhere classified |
| J13* | Pneumonia due to Streptococcus pneumoniae |
| J14* | Pneumonia due to Haemophilus influenzae |
| J15* | Bacterial pneumonia, not elsewhere classified |
| J16* | Pneumonia due to other infectious organisms, not elsewhere classified |
| J17* | Pneumonia in diseases classified elsewhere |
| J18* | Pneumonia, organism unspecified |
| J20* | Acute bronchitis |
| J21* | Acute bronchiolitis |
| J22* | Unspecified acute lower respiratory infection |
| J80* | ARDS (related to respiratory infection) |
| U071* | COVID-19, virus identified |
| U072* | COVID-19, Virus not identified |
| U04* | Severe acute respiratory syndrome [SARS] |
| **SUS OPCS-4 code list** | |
| X52* | Oxygen use |
| E85* | Ventilation support |
| E89* | Other respiratory support |

Table S16: Flow chart demonstrating the selection of data in the study
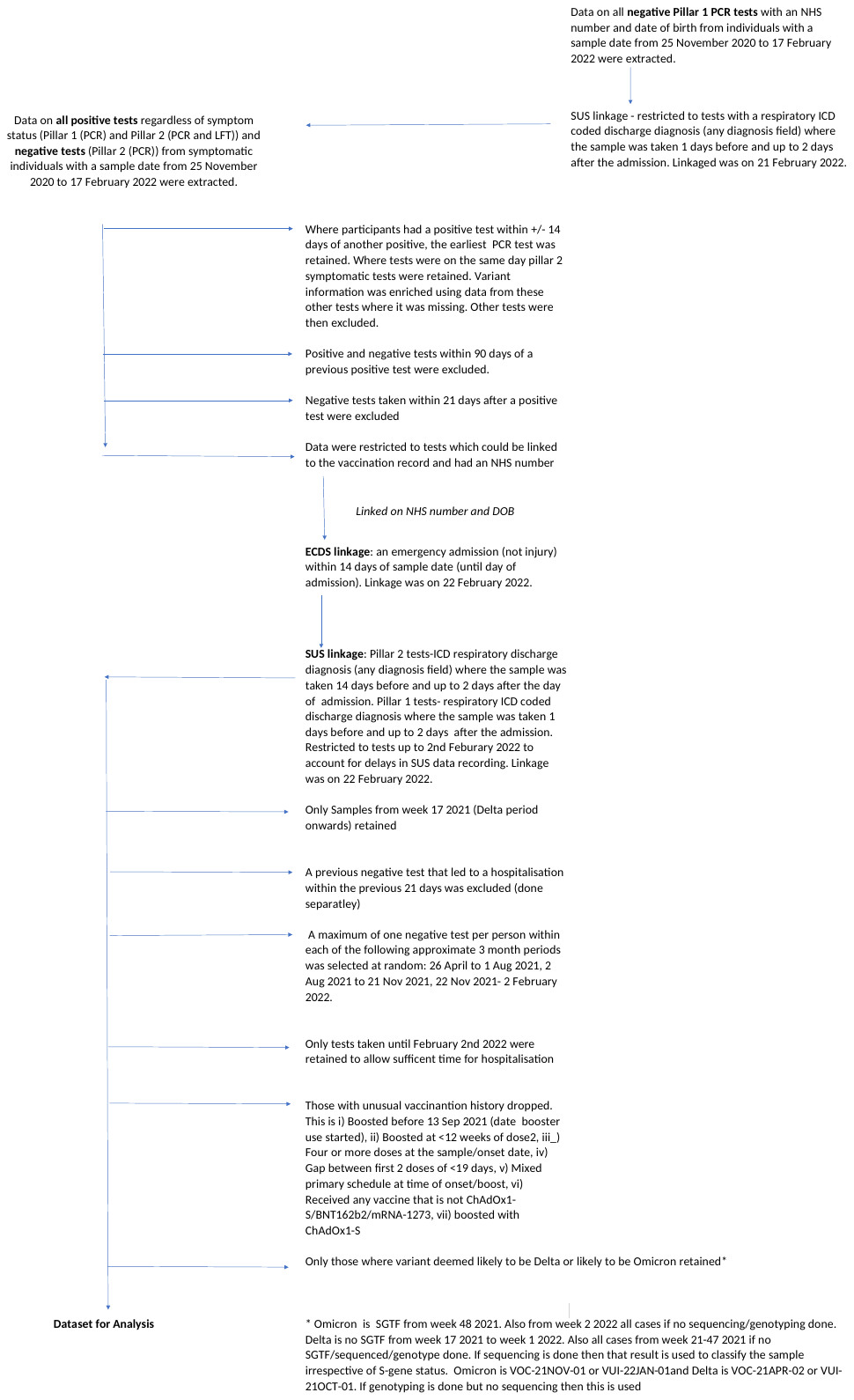
